# The Association Between Parental BMI and Offspring Adiposity: A Genetically Informed Analysis of Trios

**DOI:** 10.1101/2024.03.07.24303912

**Authors:** Liam Wright, Gemma Shireby, Tim T. Morris, Neil M. Davies, David Bann

## Abstract

**Background:** Children with obesity are more likely to have parents with obesity, too. Several environmental explanations have been proposed for this correlation, including foetal programming, and parenting practices. However, body mass index (BMI) is a highly heritable trait; child-parent correlations may reflect direct inheritance of adiposity-related genes. There is some evidence that mother’s BMI associates with offspring BMI net of direct genetic inheritance, consistent with both intrauterine and parenting effects, but this requires replication. Here we also investigate the role of father’s BMI and diet as a mediating parenting factor.

**Methods:** We used Mendelian Randomization (MR) with genetic trio (mother-father-offspring) data from 2,621 families in the Millennium Cohort Study, a UK birth cohort study of individuals born in 2001/02, to examine the association between parental BMI (kg/m^2^) and offspring birthweight and BMI and diet measured at six-time points between ages 3y and 17y. Paternal and maternal BMI were instrumented with polygenic indices (PGI) for BMI also conditioning upon offspring PGI. This allowed us to separate direct and indirect (“genetic nurture”) genetic effects. We compared these results with associations obtained using multivariable regression techniques using phenotypic BMI data only, the standard research approach.

**Results:** Mother’s and father’s BMI were positively associated with offspring BMI to similar degrees. However, in MR analysis, associations between father’s BMI and offspring BMI were close to the null, suggesting phenotypic associations reflect the direct transmission of genetic traits. In contrast, mother’s BMI was consistent in MR analysis with phenotypic associations. Maternal indirect genetic effects were between 20-50% the size of direct genetic effects. There was inconsistent evidence of associations with offspring diet. Mother’s, but not father’s, BMI was related to birthweight in both MR and multivariable regression models.

**Conclusion:** Results suggest that maternal BMI may be particularly important for offspring BMI: associations may arise due to both direct transmission of genetic effects and indirect (genetic nurture) effects. Associations between father’s and offspring adiposity that do not account for direct genetic inheritance may yield severely biased estimates of paternal influence.

## Introduction

Obesity rates among children and adolescents have risen sharply over the past five decades (Abarca-Gómez et al., 2017). This has motivated a large body of research on factors that contribute to high body mass index (BMI) among young people (Blüher, 2019; Kansra et al., 2021). One focus of this research has been on the role of parents, particularly the role of parental adiposity. Children with obesity are more likely to have parents with obesity, too (Campbell & McPherson, 2019; Heslehurst et al., 2019; Wang et al., 2017; Yu et al., 2013).

Several reasons have been given to explain this correlation. The developmental overnutrition hypothesis (DOH; Lawlor et al., 2012) posits that higher levels of maternal adiposity while the child is *in utero* can have long-term effects on offspring BMI by increasing circulating levels of pro-inflammatory cytokines, glucose, and fatty acids in the mother (Fleming et al., 2018; Godfrey et al., 2017; N. Patel et al., 2015). These are thought to increase birth weight and ‘program’ permanent changes to offspring adiposity-related physiology and behavioural traits, including changes to appetite control and metabolism (Fleming et al., 2018; Godfrey et al., 2017; Oken & Gillman, 2003; N. Patel et al., 2015; Remmers & Delemarre-van De Waal, 2011). However, the DOH cannot explain the correlation between father’s and offspring BMI. Traits related to father’s (and mother’s) adiposity, such as diet and exercise behaviour, could instead be important postnatally through their influence on the postnatal environment in which the child develops. Aspects of the family and home environment that are particularly pertinent to BMI are parental diet (which influences food availability and behaviour modelling), *food parenting practices* (active behaviours and techniques used to influence children’s food intake; C. Patel et al., 2018), and analogous influences on exercise and physical activity (Larsen et al., 2015; Lawlor et al., 2008).

While these processes may explain a correlation between parental and offspring adiposity, BMI is a highly heritable – and polygenic (Vogelezang et al., 2020; Yengo et al., 2018) – trait; heritability estimates from twin studies range 47-90% (Elks et al., 2012; Min et al., 2013). The association between parents and their children may, therefore, reflect direct transmission of genes passed on to offspring. Only a small number of studies have used genetically-informed designs that can account for the direct genetic transmission of adiposity-related genes (Bond et al., 2022; Kong et al., 2018; Lawlor et al., 2008; Richmond et al., 2017; Schnurr et al., 2020; Tubbs et al., 2020). These exploit the fact that children inherit only half of their parents’ genomes and either use polygenic indices (PGI) for BMI constructed using parental ‘non-transmitted’ alleles or adjust for parental and offspring PGIs simultaneously, thus enabling the assessment of ‘genetic nurture’ effects operating indirectly via parental traits and behaviours (Figure 1; Davies et al., 2019).

**Figure 1:**
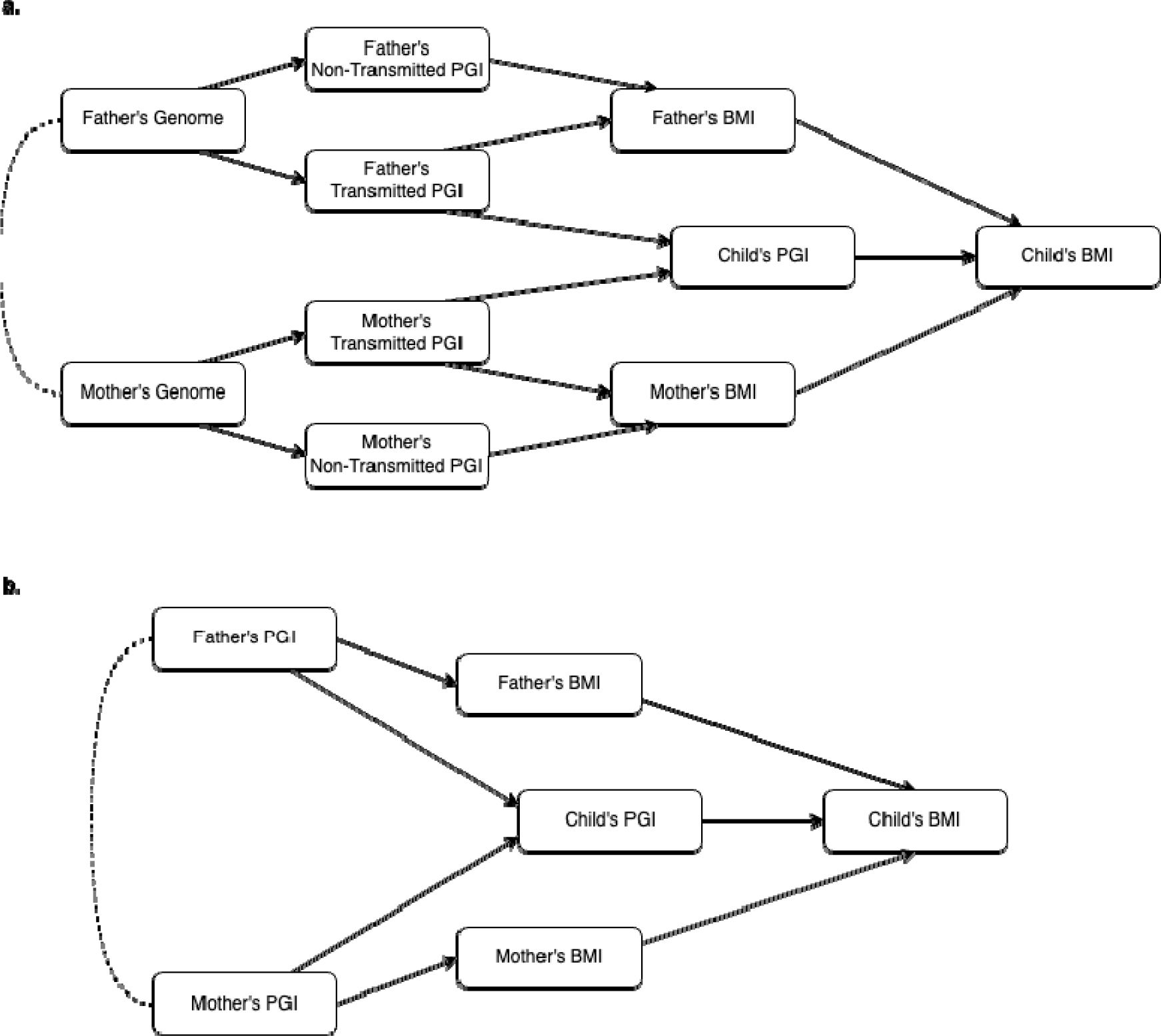
Directed Acyclic Diagram of the relationship between parental PGIs and child’s BMI. Mother’s and father’s PGI are hypothesised to be causally related to child’s BMI via transmission of genetic material to the child (i.e. through inerited alleles, as captured in the child’s PGI) and indirectly via non-transmissed alleles that influence parental BMI and consequent traits and behaviours (intrauterine environment, food parenting practices, etc.). In (a), mother’s and father’s genomes are split into transmitted and non-transmitted alleles before PGIs are calculated. In this DAG, mother’s non-transmitted PGI is a valid instrument for mother’s BMI, conditional upon the father’s transmitted and non-transmitted PGIs. Without conditioning, estimates would be biased due to assortative mating (dashed line). It is not necessary to condition upon child’s PGI to obtain unbiased estimates. In (b), PGIs are calculated using all alleles. Mother’s PGI is a valid instrument for mother’s BMI, conditional upon child’s PGI and father’s PGI. If father’s PGI is not controlled for, mother’s PGI is an invalid instrument for two reasons: first, confounding due to assortative mating (dashed line); second, collider bias as adjusting for the child’s PGI opens a path between father’s PGI and mother’s PGI. In the DAG, mother’s PGI would also be a valid instrument if father’s BMI was controlled for. However, in practice, father’s BMI is likely to be measured with error (e.g., self-report at a single time-point), and thus not would fully close the paths from mother’s PGI to child’s BMI via father’s PGI. Note, if father’s PGI does not have an indirect effect, no bias will arise.

These studies have generated inconsistent results but have differed according to the age of BMI assessment, the predictive power of PGIs used, and whether maternal and paternal genetic effects have been separated. Evidence from one regional UK birth cohort shows an association between a mother’s BMI PGI and offspring’s BMI in adolescence (Bond et al., 2022; Lawlor et al., 2008; Richmond et al., 2017; Tubbs et al., 2020). Paternal effects were not assessed, however. Studies from Denmark (Schnurr et al., 2020) and Iceland (Kong et al., 2018) found little evidence of association with parental genetics at age 18y or during adulthood, respectively – though, the former study used a relatively small sample size and the latter did not separate maternal and paternal effects. The latter two studies are also consistent with a genome-wide association study (GWAS) comparing within- and between-sibling genetic associations that found little evidence of genetic nurture influence on adult BMI (Howe et al., 2022).

The current evidence is, therefore, restricted by the partial use of genetic trio (mother-father-offspring) data, a lack of direct examination of paternal effects, and a small number of samples used. Further, there has been no investigation of the pathways through which indirect genetic effects may arise; for instance, effects upon diet, which (as noted) could explain correlations in father-child BMI. Therefore, in this study, we investigated whether maternal and paternal genetics have indirect effects upon offspring outcomes by performing an MR analysis of over 2,600 genetic trios in the Millennium Cohort Study, a UK birth cohort with repeat assessment of children’s BMI and diet from early childhood to late adolescence.

## Methods

### Participants

Participants in the Millennium Cohort Study (MCS) were born between 2000/02, decades after childhood obesity rates started to rise in the UK (Office for Health Improvement and Disparities, 2022; Stamatakis et al., 2005). Participants have been followed up on multiple occasions, beginning at 9 months and with the most recent data collection available at age 17y. (An additional sample of 4% of participants were recruited at age 3y.) Cohort member’s caregivers and, latterly, the cohort members themselves, have been interviewed at each sweep, with a wide variety of survey and biomedical data collected. At age 14y, cohort members and their resident biological parents provided saliva samples, which were used for genotyping. Further detail on the study, including genetic data collection, is available in cohort profiles (Connelly & Platt, 2014; Fitzsimons et al., 2022). Ethical review for each sweep was provided by a National Health Service Research Ethics Committee (see Shepherd & Gilbert, 2019, for more detail).

The MCS used a clustered stratified sampling design, with oversamples of children living in disadvantaged areas, from ethnic minority backgrounds, or from Scotland, Wales or Northern Ireland. We limited our analysis to singletons of (White) European ancestry/ethnicity (n = 15,456; 81.4% of the recruited sample) as the BMI PGI we used was drawn from a GWAS of European ancestry samples (Yengo et al., 2018). (For genotyped individuals, European ancestry was determined using the GenoPred pipeline [see below]; for non-genotyped individuals, self-reported White European ethnicity was used.) Only parents who were biologically related to cohort members were eligible for genetic data collection; biologically related mothers or fathers who did not reside with their child were not followed-up. As such, sample sizes for trios were limited. 2,629 families had full genetic trio data (44.8% those observed with both biological parents resident at age 14y, 17% of the total sample of European ancestry) and 5,368 had genetic data from mother-offspring pairs (59.3% of those observed with resident biological mother at age 14y, 34.7% of the total sample with European ancestry).

### Genotyping

Genotyping was carried out in the Bristol Genetics Labs (Bristol, UK) using Illumina Infinium global screening arrays-24 v1.0. For more details on the collection of samples, DNA extraction methods, and laboratory procedures, see Fitzsimons et al. (2022). Genotype calling was performed using GenomeStudio (v2.0, Illumina) and quality control was completed using PLINK1.9 and PLINK2.0 (Chang et al., 2015). Samples that could no longer be included in the sample (e.g., due to withdrawn consent) were removed prior to QC. Individuals were excluded if they had > 2% missing data, excess heterozygosity (>3 standard deviation [SD] from the mean), or X chromosome homozygosity discordant with their reported sex (females excluded with an F value > 0.2 and males with an F value < 0.8), so long as these could not be rectified using family relationships inferred using KING.

Prior to imputation, single nucleotide polymorphisms (SNPs) were excluded if they had high levels of missing data (> 3%), Hardy-Weinberg equilibrium P < 1e-6 (based on a subset of unrelated, European samples), or minor allele frequency (MAF) < 1%. The genetic data were then recoded as vcf files before uploading to the TOPMed Imputation Server, which uses Eagle2 to phase haplotypes and Minimac4 (Fuchsberger et al., 2015) with the TOPMed reference panel. Imputed genotypes were then filtered with PLINK2.0alpha, excluding SNPs with an R2 INFO score < 0.8, and recoded as hard-calls into binary PLINK format. Proceeding with PLINK1.9, samples with > 2% missing values were removed and SNPs were excluded if they had > 3% missing values, > 2 alleles or a MAF of < 1%. Duplicate samples were also removed, with the sample with the higher genotyping rate retained. In these steps, European individuals were identified using the GenoPred pipeline which involved (a) merging the MCS genotypes with data from 1000 genomes Phase 3, (b) linkage disequilibrium pruning overlapping SNPs such that no pair of SNPs within 1000 bp had r^2^ > 0.20 and (c) using an elastic net model to identify Europeans versus non-Europeans.

### Measures

#### Parental Body Mass Index

Parental height and weight were obtained via self-report at 9 months, 3y, 5y, and 7y sweeps. Weight was asked of fathers and non-pregnant mothers only, and mothers were additionally asked at 9 months for their weight pre-pregnancy. To avoid retrospective measurement error and maximise sample size, we defined BMI (kg/m^2^) for each parent using their first available, contemporary (i.e., post-pregnancy) report. Correlations between sweeps were ∼ 0.85 or greater. The majority of observations (91%) were drawn from the 9 months sweep.

#### Offspring Adiposity

Child’s height and weight were obtained at ages 3y, 5y, 7y, 11y, 14y, and 17y via direct measurement by interviewers. We converted this to BMI (kg/m^2^), which we used as our primary measure. As BMI is an imperfect measure of adiposity, particularly among children (Wang et al., 2006), we supplemented it with several other adiposity-related measures. First, as BMI changes rapidly with age during childhood and adolescence, we converted raw BMI to age and sex-adjusted z-scores using growth reference charts (the 1990 UK reference panel; Cole et al., 1998) with the `childsds` package in R (Vogel, 2020). This procedure projects observed BMI onto the distribution of BMI in the reference sample, which has been transformed to have a standard normal (mean = 0; SD = 1) distribution. Second, we used height and weight (z-scores) separately to examine whether associations were related to weight, in particular; BMI is not independent of height in children and adolescents. Third, we examined if results were similar when using direct measures of fat mass collected at ages 7y, 11y, 14y, and 17y (Staatz et al., 2021): body fat percentage, ratio of fat mass to fat free mass, fat mass index, and fat-free mass index. Fourth, we used waist-to-height ratio measured at 5y and 7y. Further detail on these variables is provided in the Supplementary Information.

Finally, to test for intrauterine effects specifically, we used birth weight (grams), collected at 9 months or 3y from parents who consulted their child’s *red book* (a portable health record), where available.

#### Offspring Diet

Child dietary data has been collected at each sweep of the MCS via reports from parents or cohort members. At ages 3y, 5y, 7y and 11y, the parent with the main responsibility for caring for the child (typically the child’s mother) was asked for the frequency or amount of consumption of various foodstuffs and drinks, with the specific questions asked changing between sweeps. At ages 14y and 17y, dietary questions were asked to cohort members instead.

From the dietary questions, we created separate variables for consumption of fruit (5y, 7y, 11y, 14y and 17y), vegetables (14y and 17y), fruit or vegetables (3y), fast food (14y and 17y), sugary drinks (11y, 14y, and 17y), and artificially sweetened drinks (11y, 14y, and 17y). Except for fruit or vegetable consumption at 3y, which is a binary variable, each of these was an ordered categorical variable with 3 or more response categories.

As self-report dietary data is captured with measurement error (Shim et al., 2014), we further used Multiple Correspondence Analysis (MCA) to extract a latent factor for each sweep capturing common variation among the individual dietary items. As only one diet question was asked at 3y, 5y and 7y, we could only perform MCA from 11y onwards. We standardized the diet factor variables (mean = 0, SD = 1) and coded all diet variables such that higher values indicated healthier diet (e.g., greater consumption of fruit and veg, lower consumption of sugary drinks and fast food, etc.). Full detail on the dietary measures, including factor loadings, is provided in the Supplementary Information.

#### Polygenic Scores for Body Mass Index

We created PGIs for mothers, fathers and children based on summary statistics from one of the largest GWAS of adulthood BMI currently available (n =∼ 700,000; sex adjusted results from Yengo et al., 2018). We constructed the PGIs using PRSice-2 (Choi & O’Reilly, 2019), disregarding ambiguous alleles and assuming additive genetic effects. To reduce the potential for horizontal pleitropy (i.e., genetic effects occurring via other traits, such as education), we used clumped genome-wide significant hits (p < 5e-8, R^2^ < 0.01, 1,000 kb window). The final PGI scores were based on 1,079 SNPs. For interpretability, we standardized the PGIs (mean = 0, standard deviation = 1) using the complete trio sample.

#### Covariates

We included variables for child’s sex and age at BMI assessment (modelled with two natural splines; Perperoglou et al., 2019), maternal age at birth, maternal years of education, family socioeconomic class (five category National Statistics Socio-economic Classification; NS-SEC), and (child’s) 10 genetic principal components (PCs). Genetic PCs were used to account for population stratification. Mother’s education and family socioeconomic class were included as potential confounders that may explain differences in child BMI and diet not directly related to parental BMI. Both were measured at age 9 months, or, if missing, at the next sweep at which data was available. Family socioeconomic class was measured as the higher of resident caregivers’ occupational class.

### Statistical Analysis

The primary aims of the analysis were to (a) examine the association between parental BMI and offspring adiposity and diet accounting for direct genetic transmission and (b) to compare this with associations obtained with the standard multivariable regression approach that does not account for genetic inheritance and relies upon phenotypic data only.

To address (a), we ran a series of multivariable Mendelian Randomization (MR) models using instrumental variables two stage least squares regression (IV-2SLS). Mother’s and father’s BMI were instrumented with parental PGIs also conditioning upon offspring PGI and the covariates listed above. Conditioning upon offspring PGI allowed us to isolate indirect from direct genetic effects (a DAG representing the analysis is shown in Figure 1b). The MR models were repeated for each outcome variable and age of assessment. We analysed each sweep separately as, based on previous results (Bond et al., 2022; Richmond et al., 2017), we anticipated effect sizes to differ by age, but had no *a priori* expectation as to the form this variation would take (e.g., linear change). For individual diet variables, we first dichotomized these and used linear probability models for the second stage regressions (see Supplementary Information for categories used for dichotomization).

To address (b), we regressed offspring adiposity and diet variables upon parental phenotypic BMI, again repeating models for each outcome variable and age of assessment – parental phenotypic BMI was not instrumented and instead entered into models directly. We again adjusted for the covariates listed above, but did not control for offspring PGI following standard practice. To compare estimates from MR and phenotypic regression models, we calculated the difference in coefficients between models, obtaining confidence intervals using bootstrapping accounting for complex survey design (Rao and Wu method, 500 bootstrap samples; Kolenikov, 2010; Rao & Wu, 1993).

As a further analysis, we estimated multivariable regression models using parental PGIs (instead of parental phenotypic BMI) directly as including mother’s, father’s and child’s PGIs in models simultaneously allowed us to assess the relative size of direct (child’s PGI) and indirect (mother’s and father’s PGIs) genetic effects upon offspring adiposity and diet.

To ensure results were not driven by outliers, we deleted values which were three or more standard deviations away from the (sweep-specific) sample mean. To maximise power, we used regression-specific complete case data. Sample sizes ranged 1,652-2,532. Sample sizes differed across analyses due to missing data for outcome variables or covariates, loss to follow-up, death, or emigration. Given this, in sensitivity analyses, we used multiply imputed data instead. As the level of selection and attrition was high (trio data available for < 20% of the eligible sample), we imputed to two samples: the set of families with genotyped trios and the set of families with genotyped duos (mother-offspring or father offspring pairs; n = 5,921; 38.3% of total European ancestry sample). We also repeated models dropping father’s PGI and using observations from genotyped mother-offspring pairs (n = 5,368; 34.7% of the total European ancestry sample) as this allowed us to double the sample size compared to the genetic trios and thus obtain more precise estimates. Further, we descriptively examined whether missingness and attrition were related to outcome variables and participant characteristics to assess the likely degree and direction of bias.

As the MCS uses a stratified survey design, we conducted analyses using sample recruitment weights and strata with the `survey` package in R (Lumley, 2004). Data cleaning and analyses were performed using R version 4.3.1 (R Core Team, 2023). The code used to run the analysis is available at https://osf.io/5vfnq/.

### Role of the Funding Source

The funders had no final role in the study design; in the collection, analysis, and interpretation of data; in the writing of the report; or in the decision to submit the paper for publication. All researchers listed as authors are independent from the funders and all final decisions about the research were taken by the investigators and were unrestricted.

## Results

### Descriptive Statistics

The mean, variation, and skewness of BMI increased as the cohort aged (Figure 2). The diet MCA factor variable was highly variable across sweeps and only weakly correlated with contemporary child BMI z-scores (−.05 < ρ < .00; Supplementary Figure S1). Descriptive statistics for the individual diet variables are shown in Supplementary Figure S2.

**Figure 2:**
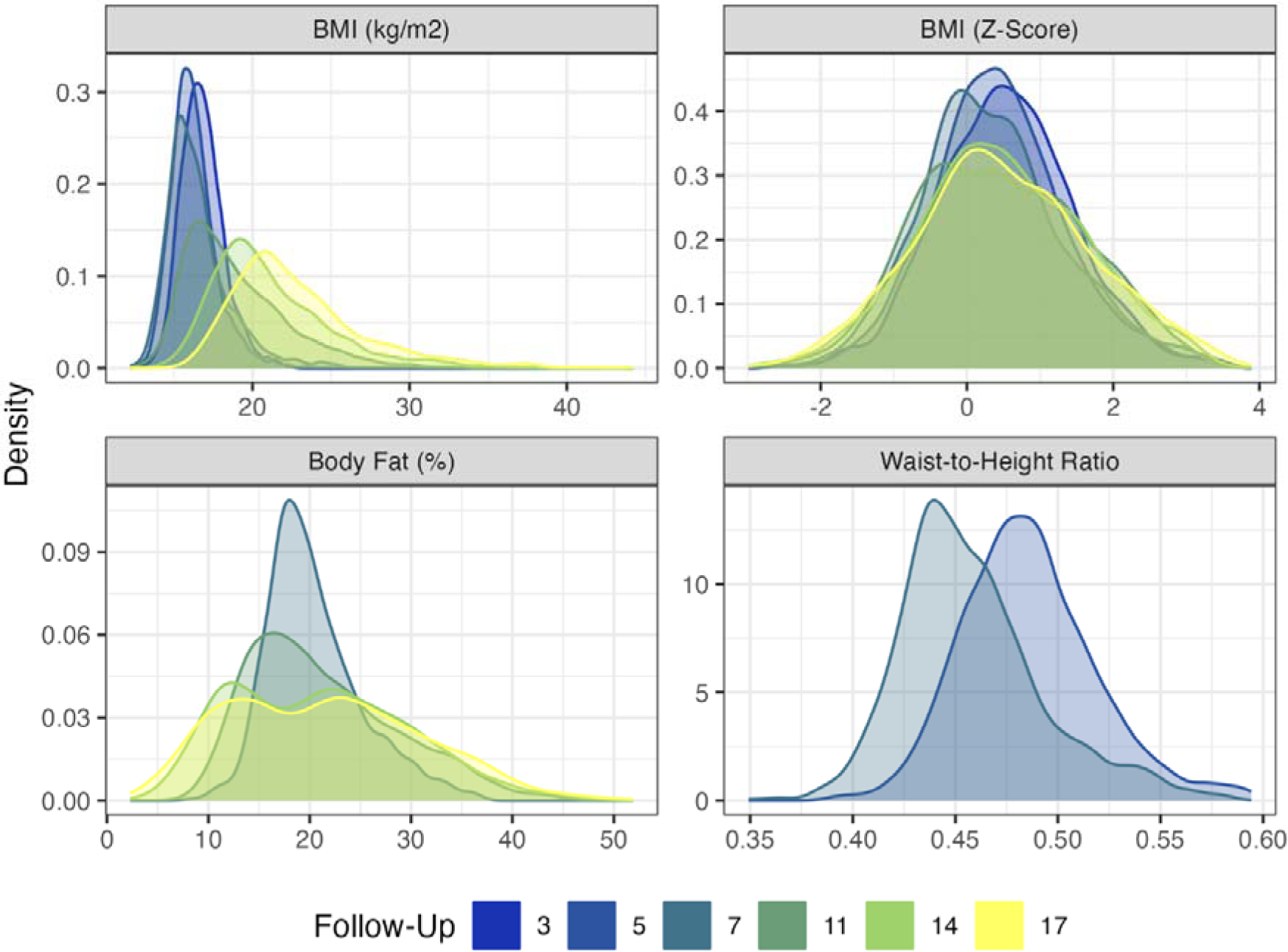
Distribution of four adiposity measures by survey sweep. Weighted with recruitment weights.

The PGIs were correlated with several measured confounders (Table 1). Higher levels of each PGI were associated with less advantaged family social class, fewer years of maternal and paternal education, and earlier maternal or maternal age at birth. However, conditioning upon parental PGIs, child’s PGI was only associated with (own) country of birth and maternal age at birth (p < 0.05). Maternal and paternal BMI were weakly correlated (ρ = .22) and the correlation between mother’s and father’s PGIs was ρ = −.02. Each parent’s PGI was also unrelated to the other parent’s BMI. Parental PGIs were strong instruments for parental BMI (F-statistics > 40 in each case).

**Table 1:** Descriptive Statistics. Association between PGI and covariates by person (child, mother or father). Genetic trio sample. For continuous covariates, figure shown is correlation between PGI and covariates. For categorical covariates, figures shown are mean (SD) of PGI in given category. Wald tests were performed to examine whether covariates were related to PGI. * p < 0.05; ** p < 0.01; *** p < 0.001.

|  |  | PGI |  |  |
| --- | --- | --- | --- | --- |
| Variable |  | Child | Mother | Father |
| Birthweight (g) |  | 0.03 | 0.05* | -0.01 |
| Sex | Male | 0.01 (1) | -0.02 (1.05) | 0.03 (1.02) |
|  | Female | 0.01 (0.98) | -0.06 (0.94) | 0.03 (1.01) |
| Maternal Age at Birth |  | -0.1*** | -0.09*** | -0.03 |
| Paternal Age at Birth |  | -0.08*** | -0.06* | -0.05* |
| Mother's BMI |  | 0.12*** | 0.25*** | 0.02 |
| Father's BMI |  | 0.13*** | 0.03 | 0.24*** |
| Family social class (NS-SEC) | Managerial and Professional | -0.04 (0.98)** | -0.09 (0.99)** | 0.01 (1.02)* |
|  | Intermediate | -0.04 (1) | -0.06 (1.06) | -0.04 (1) |
|  | Small Employer and Self-Employed | -0.03 (1.02) | -0.05 (0.95) | -0.11 (1.04) |
|  | Lower Supervisory & Technical | 0.21 (0.99) | 0.22 (1.03) | 0.19 (0.96) |
|  | Semi-Routine and Routine | 0.23 (0.99) | 0.1 (0.93) | 0.2 (1.07) |
|  | Not Working | 0.09 (0.87) | 0.13 (0.94) | 0.13 (0.95) |
| Mother's Years of Education |  | -0.08** | -0.09*** | -0.04 |
| Father's Years of Education |  | -0.08*** | -0.07** | -0.07*** |
| Country of Birth | England | 0.01 (0.99) | -0.04 (1) | 0.04 (1.02) |
|  | Wales | 0.02 (1) | -0.03 (1.04) | 0.05 (0.97) |
|  | Scotland | -0.05 (0.97) | -0.04 (0.95) | -0.03 (1.01) |
|  | Northern Ireland | 0.04 (1) | -0.04 (0.98) | -0.09 (1.03) |

### Parental BMI and Offspring Adiposity

Mother’s and father’s BMI were consistently related to higher child BMI in standard (phenotypic) multivariable regression models (Figure 3; see Supplementary Table S1 for full regression results). Associations were stronger at older ages and similar in size for mother’s and father’s BMI. A 1 kg/m^2^ increase in mother’s BMI was associated with a 0.25 kg/m^2^ (95% CI = 0.20, 0.30) increase in child’s BMI at age 17y. The corresponding figure for father’s BMI was 0.29 kg/m^2^ (0.24, 0.35).

**Figure 3:**
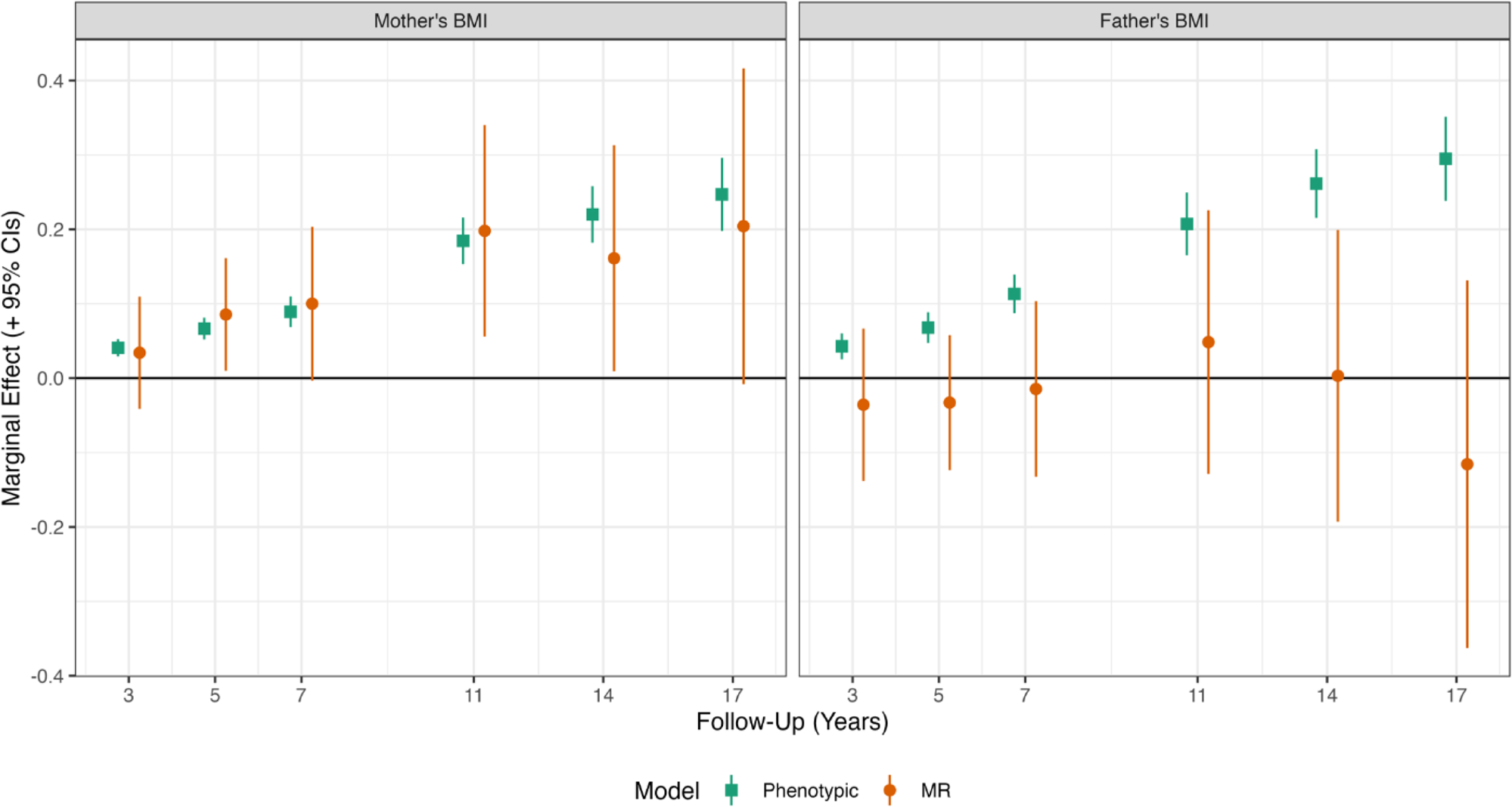
Association between mother’s and father’s BMI and offspring BMI by survey sweep. Derived from Mendelian Randomization (MR; IV 2SLS) and (phenotypic) multivariable regression of BMI on mother’s and father’s BMI, with adjustment for child’s PGI, sex, age at follow-up (two natural splines), maternal age at birth, family social class, mother’s education years, and 10 genetic principal components. In MR analysis, parental BMI instrumented with mother’s and father’s PGI. All regressions weighted with recruitment weights accounting for the cluster stratified sampling design. N = 2,130-2,468

Mother’s BMI remained consistently related to higher child BMI in MR analysis (left plot, Figure 3). Effect sizes were similar to those obtained using phenotypic multivariable regression; tests comparing coefficients across phenotypic and MR models showed no clear evidence of differences at any age. However, estimates from MR models were less precise than those from phenotypic models. In the MR model, a 1 kg/m^2^ increase in mother’s BMI was associated with a 0.20 kg/m^2^ (−0.01, 0.42) increase in child’s BMI at age 17y.

Point estimates for the association between father’s BMI and child’s BMI in MR models were small in size and typically close to the null. For instance, at age 14y, a 1 kg/m^2^ increase in father’s BMI was associated with a 0.00 kg/m^2^ (−0.19, 0.20) increase in child’s BMI. Z-tests showed clear evidence that associations obtained in MR models were attenuated relative to those using phenotypic multivariable regression (p < 0.05 from age 5y onwards).

Regarding alternate measures of adiposity, mother’s and father’s BMI were related to child’s BMI z-scores and weight z-scores but not height z-scores in multivariable regression models (Supplementary Figure S3). Again, associations for mother’s BMI from MR models were broadly consistent with those from multivariable regression models, while associations for father’s BMI were attenuated to the null. Mother’s and father’s BMI were consistently related to waist-to-height ratio, fat mass, and fat-to-fat-free ratio in phenotypic multivariable regression models but there was little consistent evidence of association in MR models, though estimates were typically imprecise (Supplementary Figure S4). There was, however, consistent evidence in MR models that mother’s BMI was related to fat-free mass index (second row, Supplementary Figure S4). This may suggest associations between mother’s BMI and child’s BMI are not driven by effects on adiposity.

Mother’s BMI was positively related to child’s birthweight in both (phenotypic) multivariable and MR models (Supplementary Table S1). The phenotypic association was 14.53 grams (9.62, 19.45) per 1 kg/m^2^ increase in mother’s BMI, while the association obtained in MR models was 23.04 grams (−0.75, 46.84) per 1 kg/m^2^. There was little evidence that father’s BMI was associated with child’s birthweight in either phenotypic or MR regression models. Corresponding figures were 1.85 grams (−3.55, 7.25) and −7.81 grams (−35.18, 19.57), respectively.

### The Association Between Parental BMI and Child Diet

Mother’s BMI was inversely related to the child diet MCA factor in (phenotypic) multivariable regression models at ages 14y and 17y, indicating a less healthy diet for children whose mother’s had higher BMI (Figure 4, left panel; see Supplementary Table S1 for full regression results). Effect sizes were small, however: a 1 kg/m^2^ increase in mother’s BMI was related to a 0.03 SD (0.01, 0.04) lower child diet factor score at age 17y. Point estimates were larger (in absolute terms) at 14y and 17y in MR than phenotypic regression models, though again effects sizes were small; the MR model predicted a 1 kg/m^2^ increase in mother’s BMI was related to a 0.05 SD (0.00, 0.10) lower child diet factor score at age 17y.

**Figure 4:**
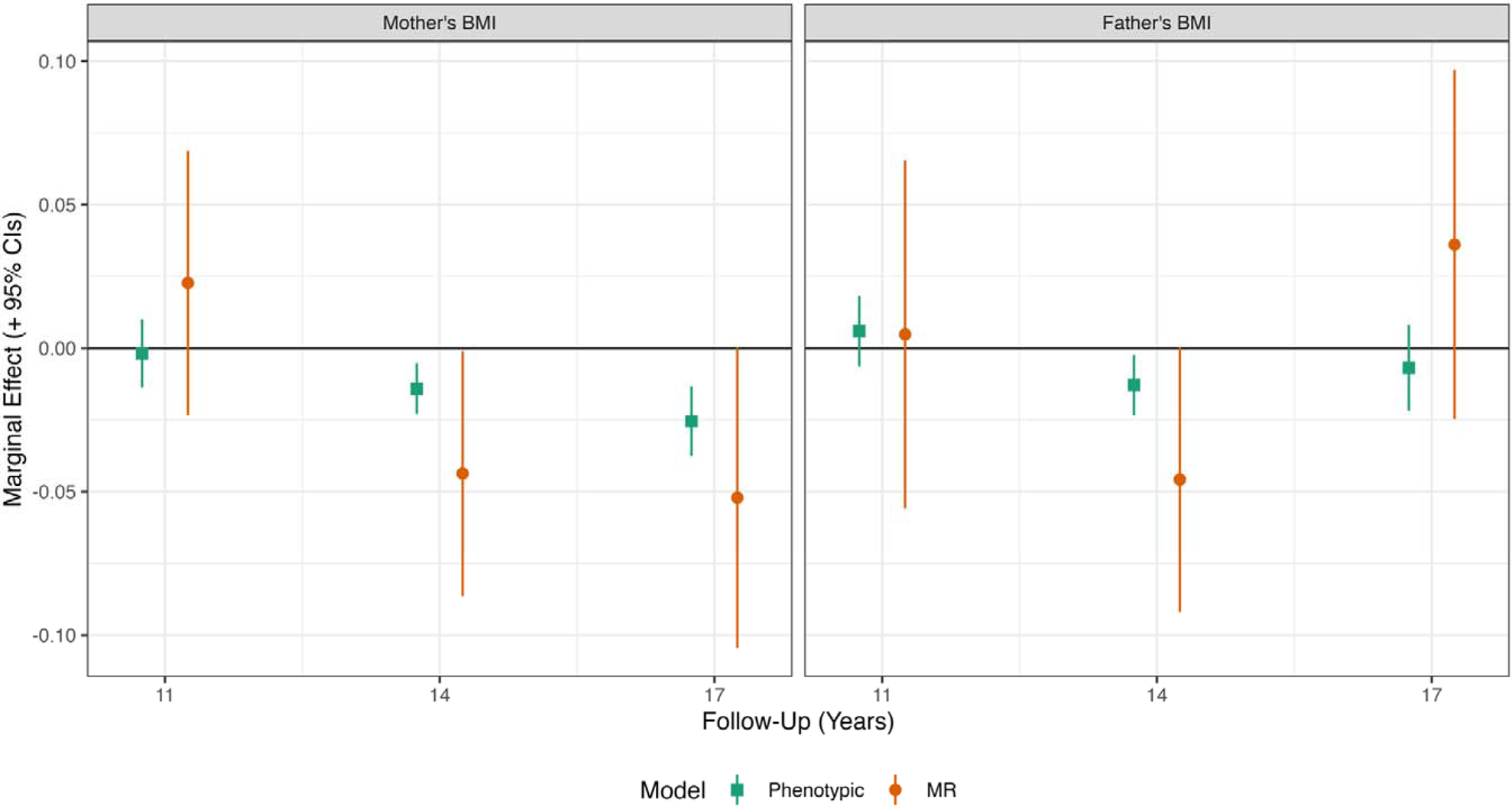
Mother’s and father’s BMI and offspring diet by survey sweep. Derived from Mendelian Rnadomization (MR; IV 2SLS) and ‘phenotypic’ multivariable regressions of offpsring on mother’s and father’s BMI, with adjustment for child’s PGI, sex, age at follow-up (two natural splines), maternal age at birth, family social class, mother’s education years, and 10 genetic principal components. In MR analysis, parental BMI instrumented with mother’s and father’s PGI. Offpsring diet captured extracting a factor from multiple diet items using MCA. All regressions weighted with recruitment weights accounting for the cluster stratified sampling design.N = 1,652 – 2,514

There was little evidence of an association between mother’s BMI and child diet factor score at age 11y in either multivariable or MR models. Father’s BMI was not consistently related to child’s diet factor scores, though there was evidence of association at age 14y in both multivariable and MR models: MR results suggested a 1 kg/m^2^ increase in father’s BMI was related to 0.05 SD (0.00, 0.09) lower diet factor score at age 14y.

Looking at individual diet items, there was evidence from MR models that mother’s BMI was related to greater frequency of fast food consumption at 14y, greater consumption of sweetened drinks at ages 11y and 14y, and lower consumption of fruit at age 11y (left column, Supplementary Figure S5). There was little clear association between father’s BMI and any individual child diet items in MR models (right column, Supplementary Figure S5).

### Comparison of Direct and Indirect Genetic Effects

In models examining associations between BMI and child’s and parent’s PGI, the (conditional) association between mother’s PGI and offspring BMI was approximately one-fifth to one-half the size of the association found for offspring’s PGI (left panel, Figure 5; see Supplementary Table S2 for full regression results). This indicated that maternal indirect genetic effects comprise a substantial component of total genetic effects for this phenotype. Conditional associations between the father’s PGI and child BMI were small at each follow-up and confidence intervals overlapped the null.

**Figure 5:**
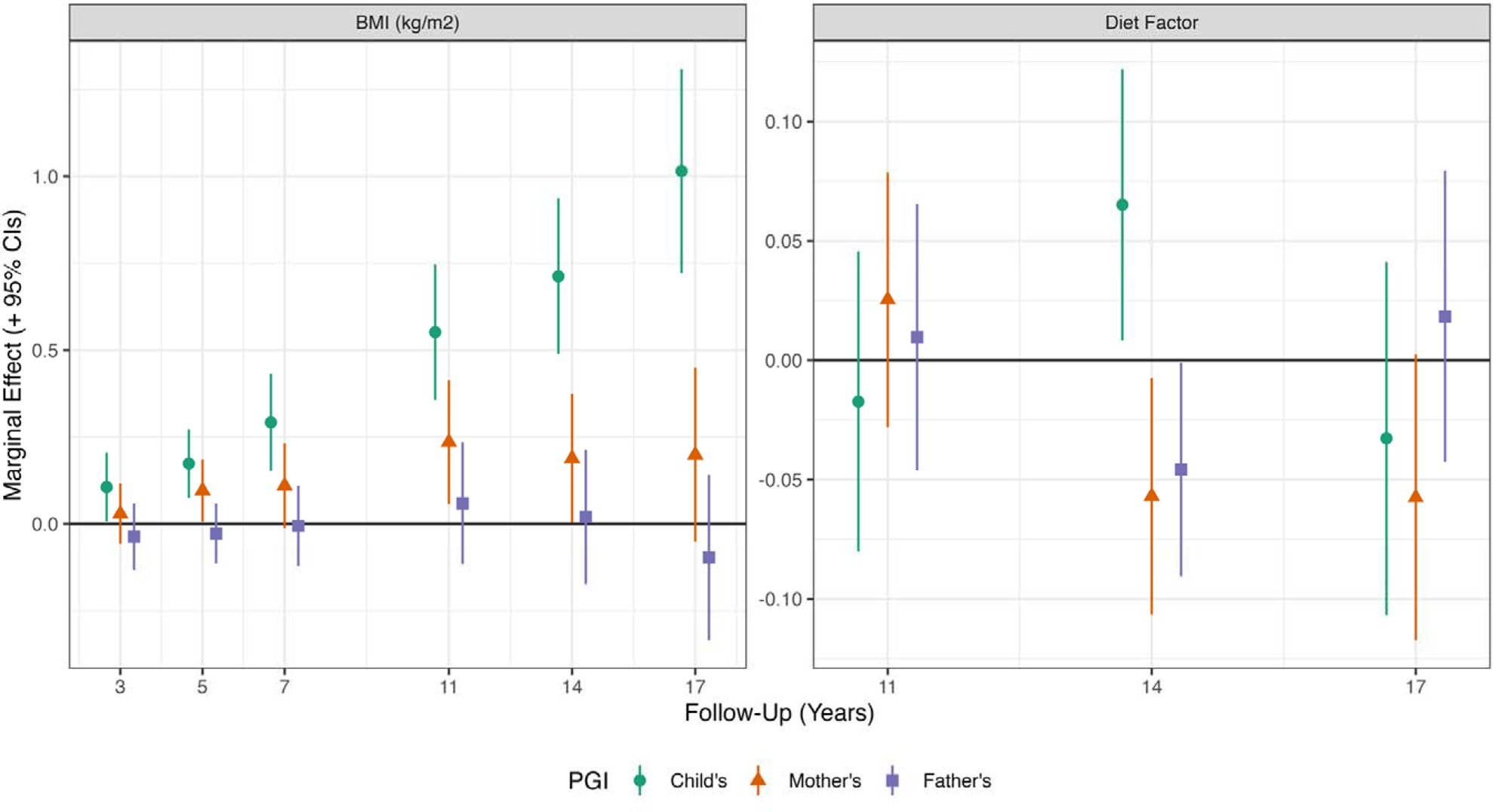
Association between parental and offspring PGI and offspring BMI z-score (left panel) and diet (right panel) by survey sweep. Derived from OLS regressions of BMI z-scores (by sweep) on mother’s, father’s and child’s adulthood PGI, with adjustment also for sex, age (two natural splines), maternal age at birth, family socioeconomic class, mother’s education year, and 10 genetic principal components. Regressions weighted with recruitment weights accounting for the cluster stratified sampling design.

The association between the mother’s PGI and birthweight (25.63 g, 95% CI = −2.49, 53.76, per 1 SD increase in the mother’s PGI) was larger than the corresponding direct genetic effect (9.26 g, 95% CI = −22.89, 41.41), though confidence intervals overlapped. Further, there was little evidence of an association between father’s PGI and birthweight (−5.55 g, 95% CI = −30.82, 19.72, per 1 SD increase in father’s PGI)

For child’s diet, mother’s PGI was associated more strongly with less healthy offspring diet than the child’s PGI was (right panel, Figure 5), though confidence intervals overlapped, except at age 14y. There was no clear or consistent association between child’s PGI and diet. In fact, at age 14y, offspring’s PGI was related to a *more healthy* diet – a 1 SD in offspring’s PGI was associated with 0.07 SD (0.01, 0.12) increase in the diet factor variable.

### Sensitivity Analyses

MR estimates were qualitatively similar when not including father’s PGI and BMI in models, both within the genetic-trio sample (middle vertical panel, Supplementary Figure S6) and in the larger sample of genotyped mother-offspring pairs (right panel, Supplementary Figure S6). MR estimates in the mother-offspring pairs were typically smaller in size, however (albeit to a limited extent). Further, in the mother-offspring pair sample, confidence intervals for the association between maternal BMI and offspring diet overlapped the null (bottom right hand panel, Supplementary Figure S6).

MR estimates were qualitatively similar for both maternal and paternal BMI when using multiple imputation (Supplementary Figures S7-S8). Point estimates for the effect of maternal BMI upon offspring BMI, diet and birthweight were smaller in the sample with genotyped mother-offspring or father-offspring pairs than in the genetic trio sample, though confidence intervals overlapped.

### Assessment of Sample Bias

Compared to eligible participants without genetic trio data, those with genetic trio data were more likely to have healthy diets, higher birthweight, an older mother, parents with greater BMI, and a more advantaged socioeconomic background (Supplementary Table S3, Column 2). There was less bias according to these factors among the sample of genotyped mother-offspring pairs, though differences remained (Supplementary Table S3, Column 3). Higher BMI z-scores were related to a lower chance of participation at subsequent sweeps (Supplementary Figure S9). For instance, a 1 SD higher BMI z-score at age 11y was related to ∼ 3.25% points lower probability of participating at 17y.

## Discussion

Using lifecourse data from over 2,600 genetic trios in the Millennium Cohort Study, we found evidence that mother’s and father’s BMI were associated longitudinally with offspring adiposity between ages 3y to 17y. Associations increased as the children aged. However, our results suggested that the association between father’s and child’s BMI was explained by direct genetic inheritance: there was little evidence of an association in MR models that accounted for transmission of adiposity related-genetic variants. Mother’s BMI was associated with offspring BMI in MR models, consistent with the effect of maternal genes operating partly indirectly via maternal traits. Indirect genetic effects were estimated to be between 20-50% of direct genetic effects for offspring BMI. However, alternate measures of adiposity, such as fat mass and waist-to-height ratio, did not show consistent positive associations with maternal BMI when accounting for genetic inheritance, which may suggest association with offspring BMI are not driven by effects on adipose tissue. There was evidence of an association between mother’s BMI and child’s diet, at least at ages 14y and 17y, and again when direct genetic inheritance was accounted for.

Our results are (partly) consistent with analyses of a regional UK birth cohort which find associations between maternal adiposity and offspring BMI in adolescence (Bond et al., 2022; Tubbs et al., 2020). Our results differ in finding clear evidence of association in earlier childhood. Further, effect sizes (when converted to SD units) were typically larger (by 50% or more) in the present study. If causal, our results suggest that intervening to reduce maternal BMI while the child is in utero could have modest long-term intergenerational effects upon obesity rates.

To our knowledge, our analysis of father’s BMI using MR is novel in the literature. The finding that father’s BMI is not associated with offspring BMI after accounting for direct genetic inheritance may explain the small effect sizes found by Kong et al. (2018) who did not separate maternal and paternal effects. However, their study also differed in measuring offspring BMI in young adulthood – parental effects could be larger in childhood and adolescence when individuals are more likely to live with their parents. Results require replication in an older sample.

Our finding that father’s BMI is unlikely to affect offspring BMI (conditional on genotype) is intriguing in light of the large number of studies investigating father-child correlations in phenotypic BMI (Campbell & McPherson, 2019; Patro et al., 2013; Wang et al., 2017). If the present results are replicated, they suggest phenotypic correlations between father’s BMI and offspring’s BMI could be explained by direct genetic inheritance and genetic confounding. Future observational studies that aim to test for environmental causes of the father-child adiposity correlations should adopt genetically informed designs (such as, the adoption study). However, it is possible that paternal indirect genetic effects may be stronger in contexts where fathers are more involved in childcare – future studies should also examine genetic nurture in countries marked by greater gender equality.

In contrast to fathers, we found little evidence that direct genetic inheritance biased phenotypic estiamtes of maternal effects on offspring adiposity. Our results were little changed when we included or omitted father’s PGI and parental PGIs displayed little correlation. This is promising given that, for data availability reasons, there are typically many more mother-child pairs than full trios in available genetic studies (e.g., due to study design, cost constraints, or parental separation).

Our results are consistent with the developmental overnutrition hypothesis, in line with other MR studies (Bond et al., 2022; Tyrrell et al., 2016); mother’s BMI was also related to offspring birth weight. However, in MR models, there was also evidence that both mother’s and father’s BMI was related to offspring diet, at least at some ages, which is also consistent with a postnatal pathway. Nevertheless, dietary data were imprecisely measured – we used few measures at each sweep and these were captured in a high level self-reported way. Parental practices may have influenced offspring diet in subtle ways which are challenging to capture in population studies (e.g., relatively small increases in total calories or fat content which cumulatively impact offspring weight gain; Government Office for Science, 2007). The diet MCA factor variable also had little correlation with observed BMI. Future studies should use more robust measures of diet as well as examine other potential pathways, such as physical activity.

Future studies should also examine the role of other genetically-influenced parental traits. For instance, father’s education or cognitive ability could exhibit genetic nurture effects on offspring adiposity; though Kong et al. (2018) find no evidence for genetic nurture effects of parental BMI, they do observe genetic nurture effects on offspring BMI when using a PGI for educational attainment, specifically. The role of other traits may be investigated using a similar MR approach to that used here but could also involve GWAS of BMI that separate direct and indirect genetic effects, either through explicit study design (e.g., GWAS of trios) or through summary data methods such as genomicSEM (Warrington et al., 2021).

### Strengths and Limitations

This study had a number of strengths. We used a PGI based on data from a large GWAS of adult BMI (Yengo et al., 2018). Previous null results may be due to less powerful PGIs used (Bond et al., 2022). We used data on complete genotyped parent-offspring trio data which allowed us to better account for bias due to assortative mating and to compare maternal and paternal indirect genetic effects. Further, our data were drawn from a nationally representative cohort with multiple measurements of offspring diet and other adiposity-related traits. Data collection was also relatively recent and, as such, particularly relevant for the current obesity epidemic.

However, our sample was selected, with genetic trio data only available for 45% of those observed with resident parents at age 14y, reflecting the practical challenges of obtaining genetic data in nationally representative samples of families. Biological parental figures not present in the household were also not included. However, the target population for this analysis is arguably those parental figures who are closely involved in offspring development and growth. It should also be noted that the sample is likely more representative than several other datasets used widely in the genetics literature (e.g., UK Biobank).

There was evidence of attrition bias, with high levels of BMI related to a greater likelihood of dropping out of the study. This may have attenuated associations towards the null. While our analytical design allowed us to account for genetic inheritance, MR estimates of parental effects can still be biased by other factors, such as horizontal pleiotropy and confounding by social or familial factors. Notably, parental BMI PGIs were related to family socioeconomic position. Nevertheless, confounding factors are likely to have biased both maternal and paternal indirect genetic effect estimates and are unlikely to explain the differences in effect estimates between parents. This logic also applies to bias arising from a selected sample.

While offspring BMI and adiposity were measured objectively, diet was measured with only a few high-level self-report survey items. These did not appear to have optimal psychometrics: diet was very weakly related to BMI and the correlation between diet variables across sweeps was low. A further limitation was the measurement of parental adiposity, which was based on self-reported BMI and was captured postnatally. Self-reported BMI is typically underestimated (Gorber et al., 2007) and BMI does not distinguish fat and lean mass. This may have biased results for fathers, in particular, as BMI is less correlated with fat mass among males (Deurenberg et al., 1991; Gallagher et al., 1996). Future work should repeat the analysis using direct measures of parental adiposity. A final limitation was that we included the results of many regression models. Some results were inconsistent across ages and may be explained by sample variation.

### Conclusions

Associations between mother’s BMI and offspring BMI were robust to accounting for direct genetic transmission and consistent with mother’s BMI have a causal effect on offspring weight. Intervening to reduce mother’s BMI may have intergenerational effects on adiposity. Assocation between father’s BMI and offspring BMI were fully attenuated when accounting for direct genetic transmission. The well replicated (phenotypic) correlation between fathers and child’s BMI may reflect strong genetic confounding.

## Statements

### Declaration of Interest

All authors declare no conflicts of interest.

### Funding

The funders had no final role in the study design; in the collection, analysis, and interpretation of data; in the writing of the report; or in the decision to submit the paper for publication. All researchers listed as authors are independent from the funders and all final decisions about the research were taken by the investigators and were unrestricted. DB and LW are supported by the Medical Research Council (MR/V002147/1); DB, GS, TM and LW by the Economic and Social Research Council (ES/M001660/1). NMD is supported by a Norwegian Research Council Grant number 295989.

### Author Contributions

All authors contributed to and agreed upon the analysis plan. GS quality controlled and prepared the raw genetic data files. LW carried out the analysis and wrote the first draft of the manuscript. All author provided critical revisions and read and approved the final manuscript.

### Data Availability

Genetic data from the Millennium Cohort Study are available via application to the Centre for Longitudinal Studies, University College London (https://cls.ucl.ac.uk). The code used to run the analysis is available at https://osf.io/5vfnq/.

## Notes

### Competing Interest Statement

The authors have declared no competing interest.

### Author Declarations

Ethical approval for the Millennium Cohort Study was provided by the South West National Health Services Research Ethics Committee (MREC/01/6/19)

