## Supplementary Information for "The Association Between Parental BMI and Offspring Adiposity: A Genetically Informed Analysis of Trios"

### Measures

#### Adiposity

BMI is an imperfect measure of adiposity, particularly among children (Wang et al., 2006). It is intended to be a measure of weight that is independent of height, but this is not case when using the typical formula for BMI (kg/m^2^) in childhood. Thus, we supplemented our analysis with four measures of adiposity collected at ages 7y, 11y, 14y, and 17y following Staatz et al. (2021): body fat percentage, ratio of fat mass to fat free mass, fat mass index, and fat free mass index. Body fat percentage was measured through foot-to-foot bio-electrical impedance analysis using Tanita (Bf-522W) scales by interviewers adhering to standardised protocols. Fat mass (FM) was calculated by multiplying body mass percentage by weight (kg), and fat-free mass (FFM) was calculated as the remainder. To create fat mass and fat-free mass indices, we divided each by height (m^B^) raised to a power (B) such that the resulting indices were independent of height. We calculated B by regressing log FM or log FFM on log height and extracting the coefficient, performing these regressions for each sex and sweep separately. These coefficients are shown in the table below.

| Variable | Sex | 7y | 11y | 14y | 17y |
| --- | --- | --- | --- | --- | --- |
| Fat Mass | Male | 3.98 | 4.76 | 3.00 | 1.95 |
|  | Female | 4.32 | 4.57 | 2.91 | 3.34 |
| Lean Mass | Male | 2.43 | 2.63 | 2.59 | 2.07 |
|  | Female | 2.43 | 2.57 | 2.21 | 1.41 |

#### Diet

The individual diet questions, including sweeps collected at, response categories, and corresponding variables from the original dataset, are provided in Supplementary Table S4. We recoded items such that higher values indicated a healthier diet. The correlations between the MCA factors variables and the individual diet measures are displayed below.

| Variable | 11y | 14y | 17y |
| --- | --- | --- | --- |
| Freq. Sugary Drinks | -0.07 | 0.65 | 0.65 |
| Freq. Sweetened Drinks | 0.20 | 0.35 | 0.44 |
| Portions Fruit per Day | 0.26 |  |  |
| Freq. Fast Food |  | 0.66 | 0.66 |
| Freq. Fruit |  | 0.65 | 0.67 |
| Freq. Vegetables |  | 0.67 | 0.68 |

Individual diet items were dichotomised in regression models (full categories were used to derive diet MCA factors). The categories used are displayed in Supplementary Figures S2 and S4.

### Figures

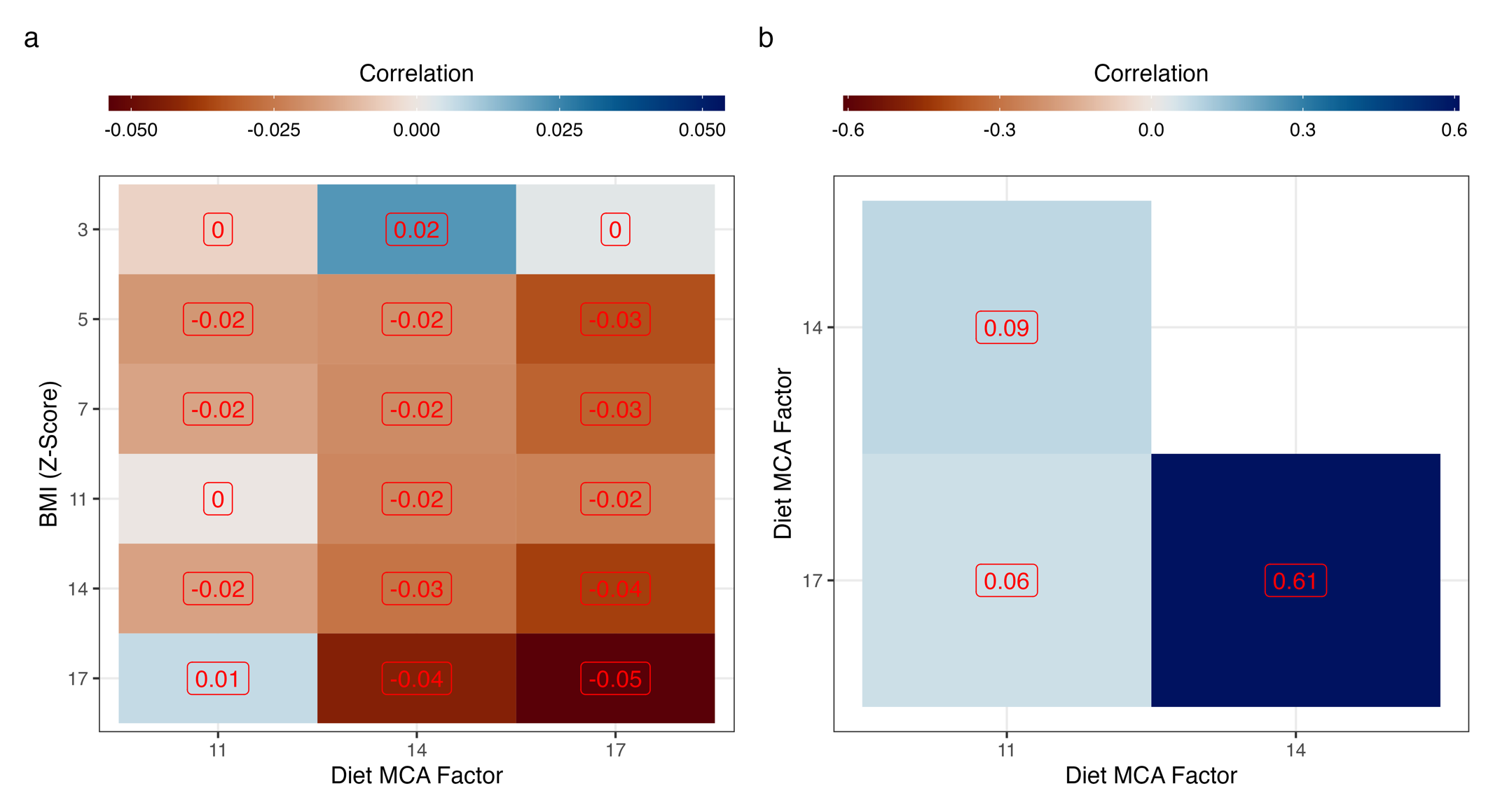

**Figure S1: Correlation between diet MCA factor and (a) BMI z-scores and (b) the diet MCA factor itself, by age at follow-up.** Diet MCA factor is coded such that higher values indicate a *healthier* diet. Correlations weighted with recruitment weights and accounting for the cluster stratified sampling design.

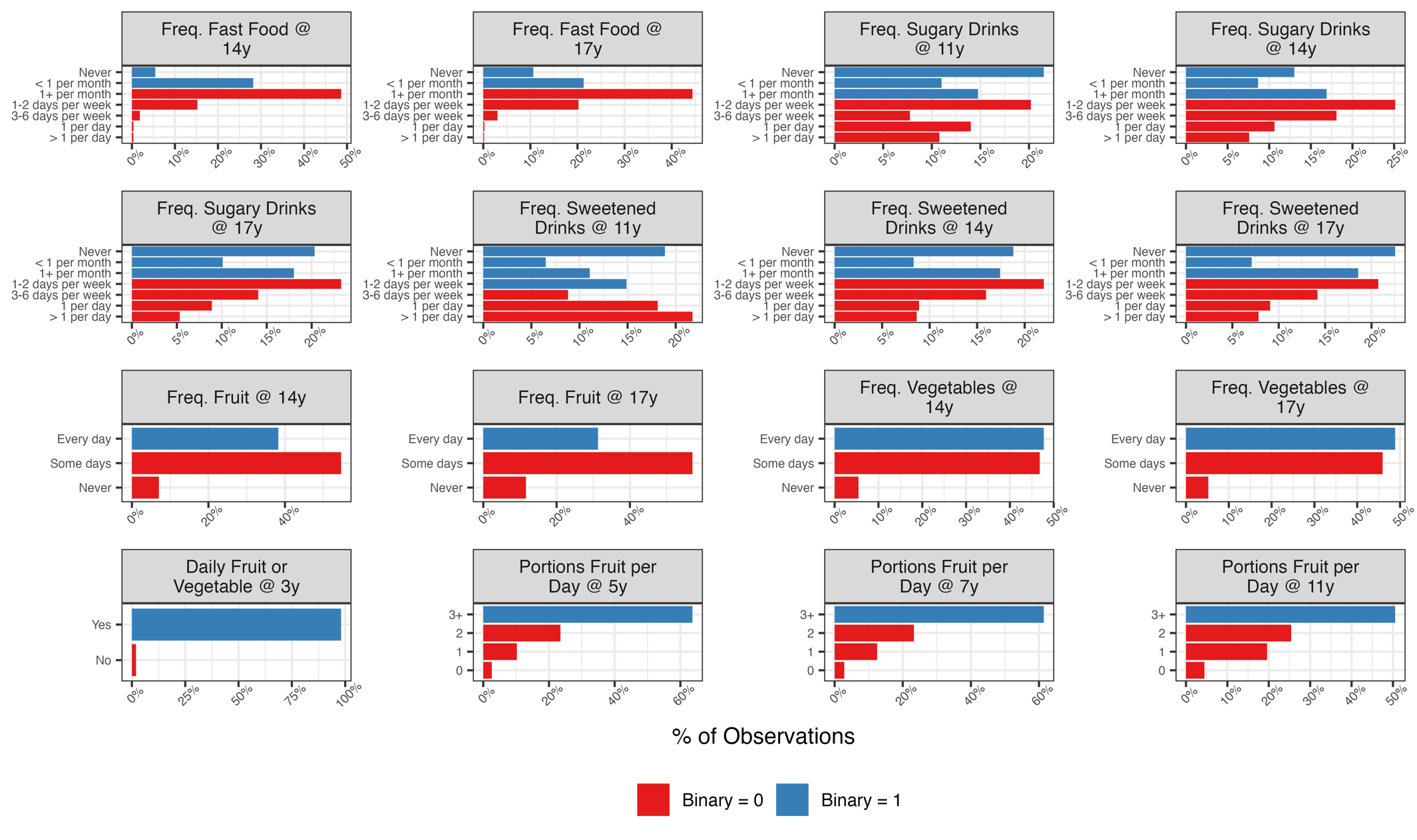

**Figure S2: Distribution of individual diet items.** Diet items were dichotomized in regression models to simplify interpretation. The colour of the bar reflects the level the category was dichotomised into (blue = 1, red = 0).

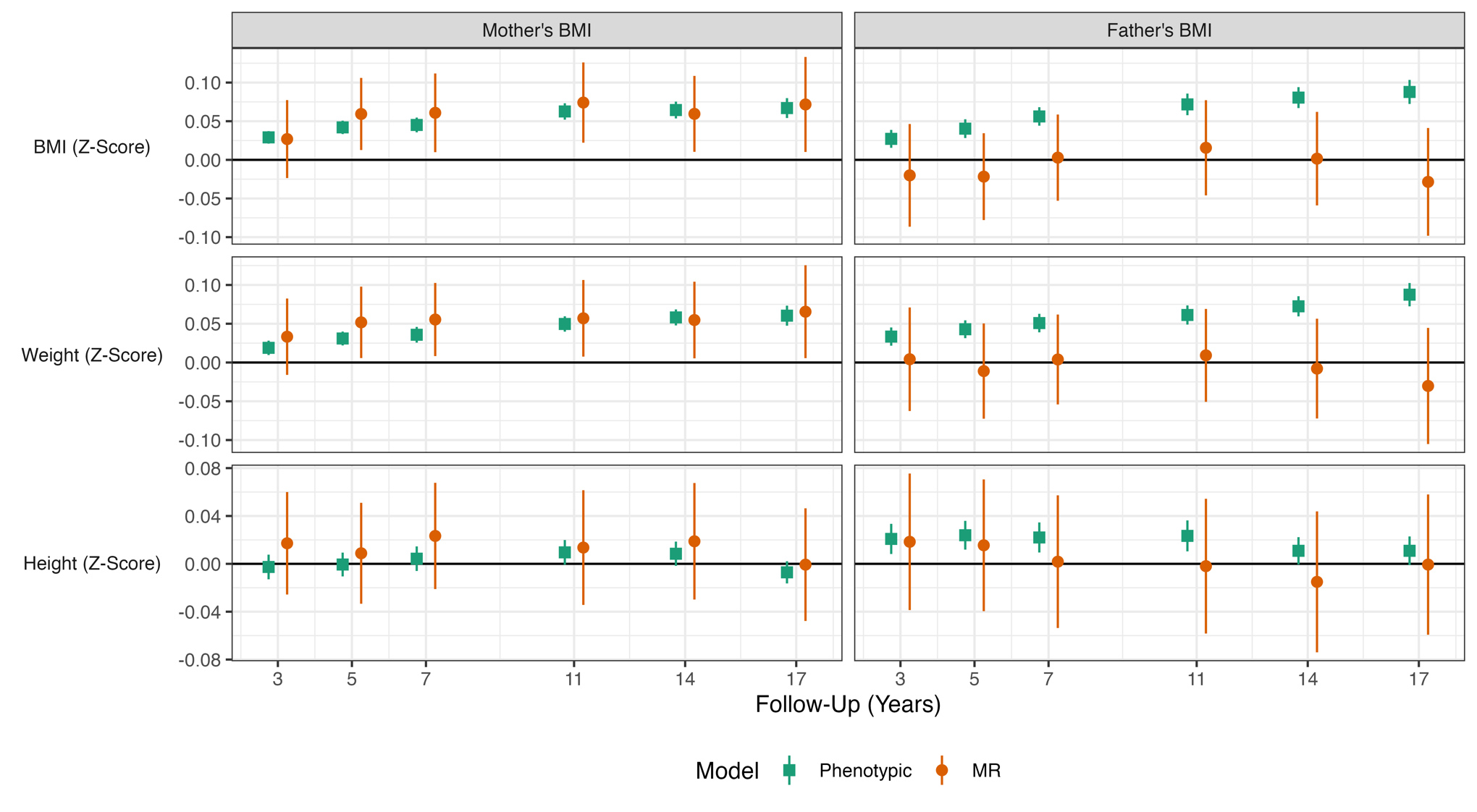

**Figure S3: Association between mother’s and father’s BMI and offspring BMI, weight, and height (z-scores) by survey sweep.** Derived from Mendelian Randomization (MR; IV 2SLS) and phenotypic multivariable regressions of offspring BMI, weight, and height (z-scores) on mother’s and father’s BMI, with adjustment for child’s PGI, sex, age at follow-up (two natural splines), maternal age at birth, family social class, mother’s education years, and 10 genetic principal components. In MR analysis, parental BMI was instrumented with mother’s and father’s PGI. All regressions were weighted with recruitment weights accounting for the cluster stratified sampling design.

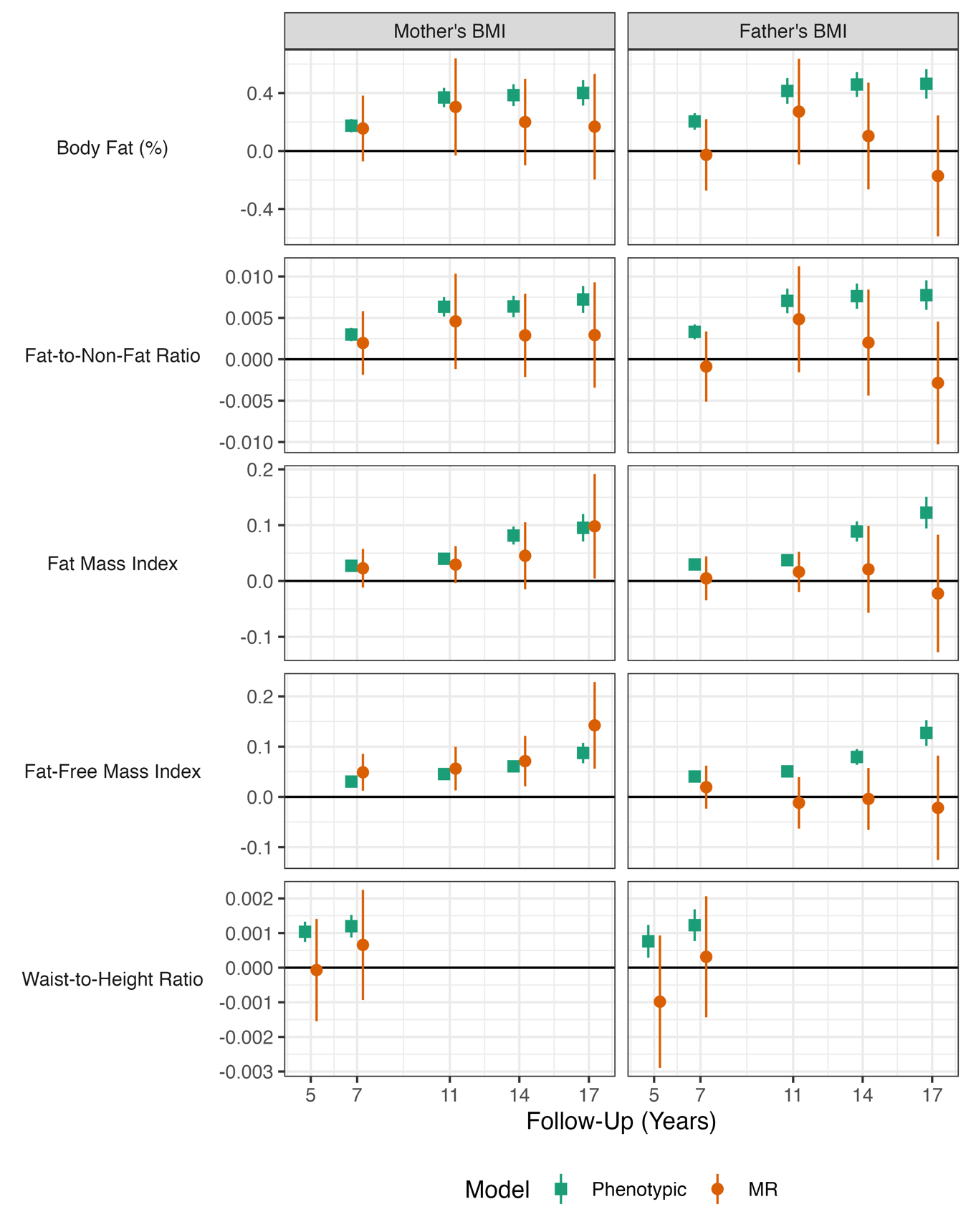

**Figure S4: Association between mother’s and father’s BMI and offspring adiposity by survey sweep.** Derived from Mendelian Randomization (MR; IV 2SLS) and phenotypic multivariable regressions of offspring adiposity on mother’s and father’s BMI, with adjustment for child’s PGI, sex, age at follow-up (two natural splines), maternal age at birth, family social class, mother’s education years, and 10 genetic principal components. In MR analysis, parental BMI was instrumented with mother’s and father’s PGI. All regressions were weighted with recruitment weights accounting for the cluster stratified sampling design.

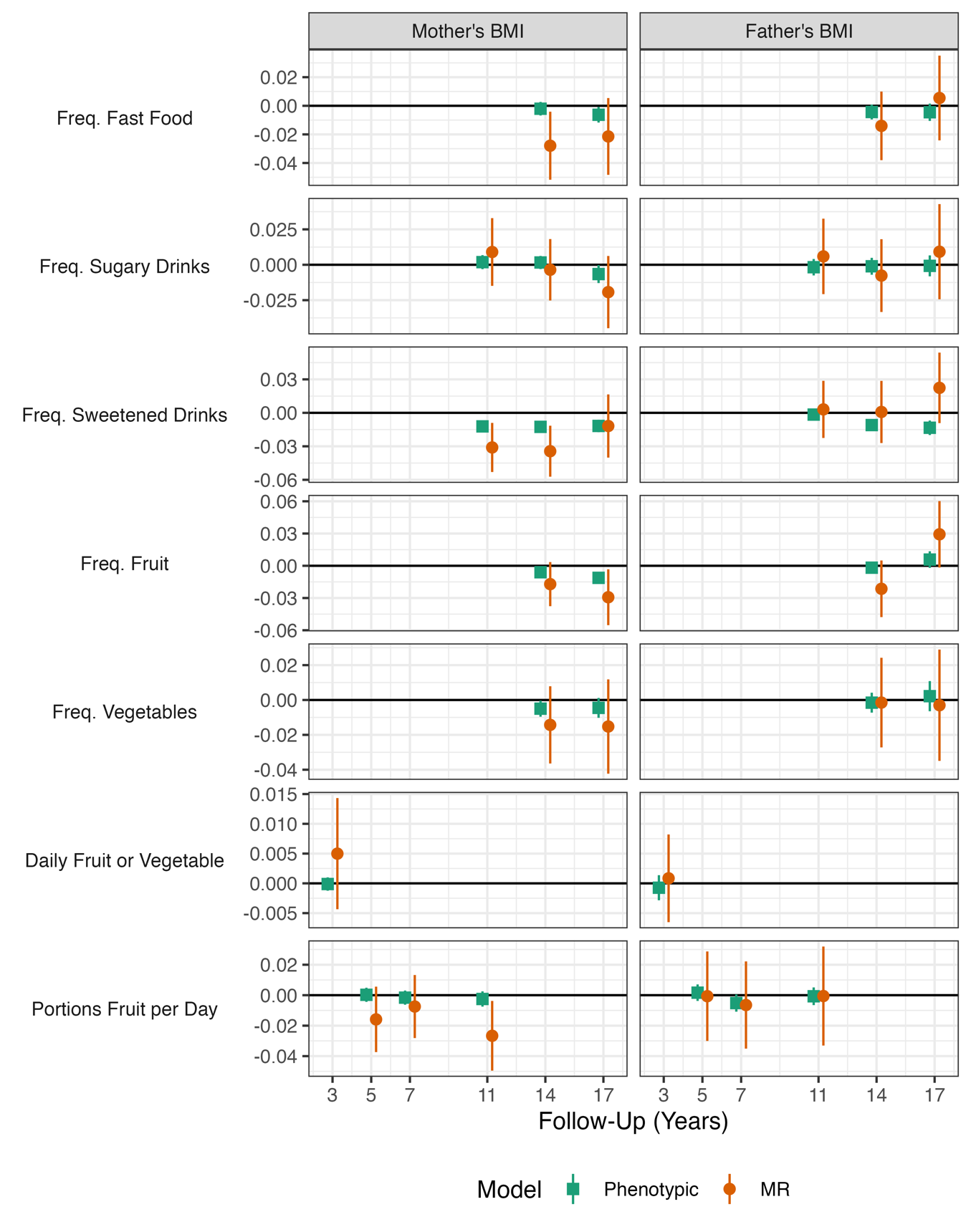

**Figure S5: Association between mother’s and father’s BMI and offspring diet by survey sweep.** Derived from Mendelian Randomization (MR; IV 2SLS) and phenotypic multivariable regressions of offspring diet on mother’s and father’s BMI, with adjustment for child’s PGI, sex, age at follow-up (two natural splines), maternal age at birth, family social class, mother’s education years, and 10 genetic principal components. In MR analysis, parental BMI instrumented with mother’s and father’s PGI. Diet variables are dichotomized (see Supplementary Figure S2 for categorisation) and coded such that a value of 1 indicated a healthier diet (associations thus represent differences in probability). All regressions weighted with recruitment weights accounting for the cluster stratified sampling design.

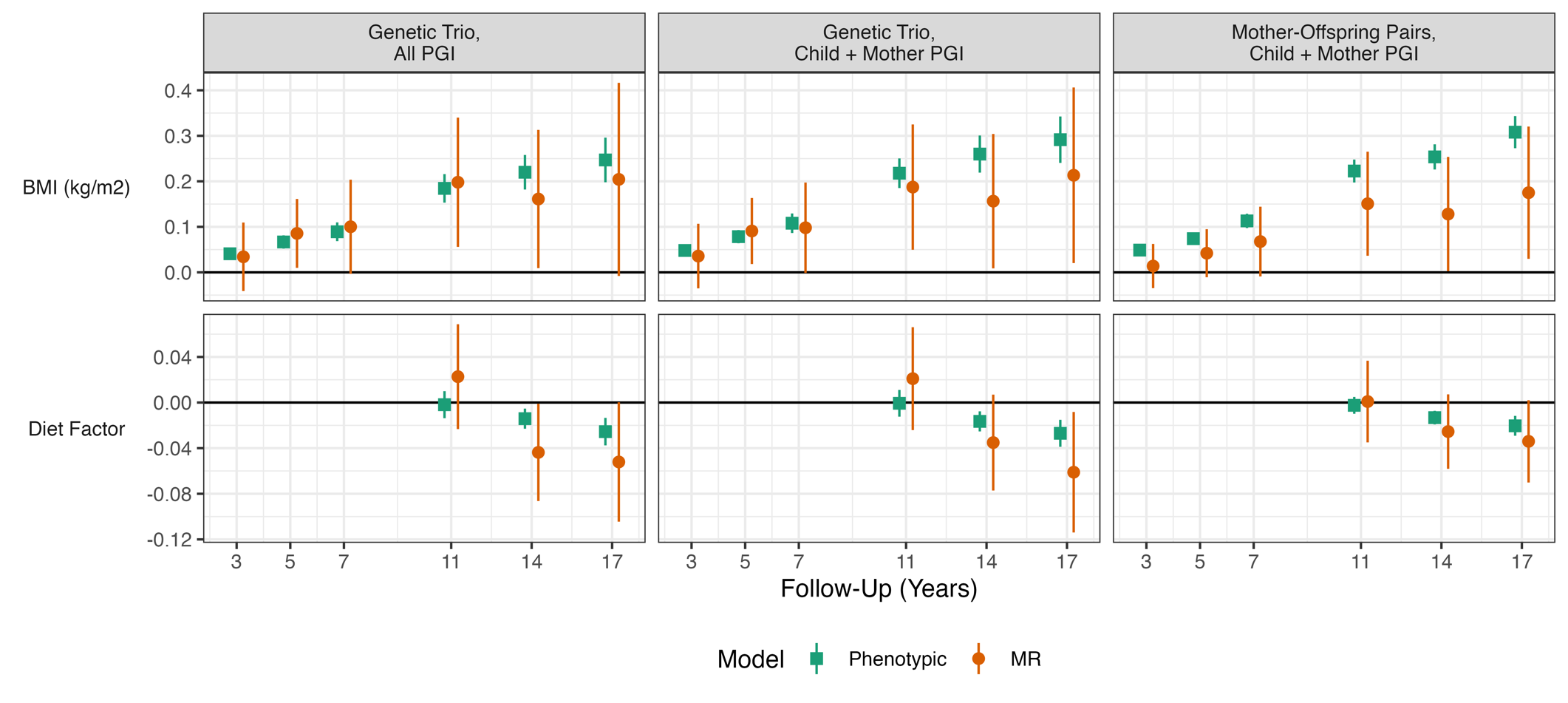

**Figure S6: Association between mother’s BMI and offspring BMI and diet by sample, PGIs used, and survey sweep.** Derived from Mendelian Rnadomization (MR; IV 2SLS) and ‘phenotypic’ multivariable regressions of offspring adiposity on mother’s BMI, with adjustment for child’s PGI, sex, age at follow-up (two natural splines), maternal age at birth, family social class, mother’s education years, and 10 genetic principal components. In MR analysis, mother’s BMI instrumented with mother’s PGI. ‘Genetic Trio, All PGI’ refers to models using genetic trio sample and including father’s BMI as a covariate and, in MR analysis, father’s PGI as an instrument for this. ‘Genetic Trio, Child + Mother PGI’ refers to models of the same sample except father’s BMI and father’s PGI was not included. ‘Mother-Offspring Pairs, Child + Mother PGI’ refers to models using the mother-offspring pair genotyped sample, with father’s PGI or BMI again not included in as covariates or instruments. This regression doubled the sample size. All models also included adjustment for sex, age (two natural splines), maternal age at birth, family socioeconomic class, mother’s education years, and 10 genetic principal components. Regressions were weighted with recruitment weights accounting for the cluster stratified sampling design.

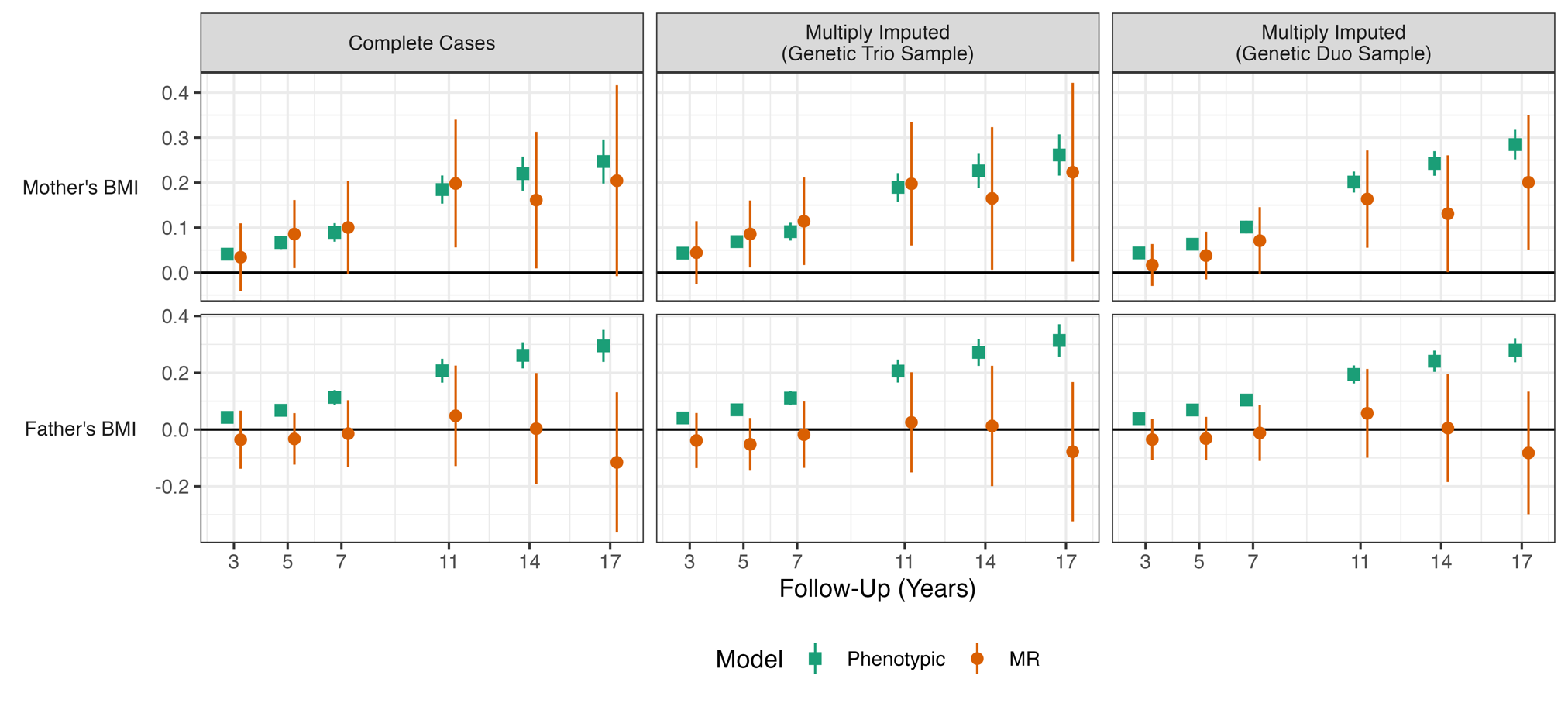

**Figure S7: Association between mother’s and father’s BMI and offspring BMI (z-scores) by sample, PGIs used, and survey sweep.** Derived from Mendelian Rnadomization (MR; IV 2SLS) and ‘phenotypic’ multivariable regressions of offspring BMI (z-score) on mother’s BMI, with adjustment for child’s PGI, sex, age at follow-up (two natural splines), maternal age at birth, family social class, mother’s education years, and 10 genetic principal components. In MR analysis, parental BMI instrumented with mother’s and father’s PGI. ‘Complete Cases’ refers to models using (outcome-sweep specific) complete cases data. ‘Multiply Imputed (Genetic Trio Sample)’ refers to models using multiply imputed data for the genotyped trio (mother-father-offspring) sample. ‘Multiply Imputed (Genetic Dup Sample)’ refers to models using multiply imputed data for the genotyped duo (mother-offspring or father-offspring) sample. All models also included adjustment for sex, age (two natural splines), maternal age at birth, family socioeconomic class, mother’s education years, and 10 genetic principal components. Regressions were weighted with recruitment weights accounting for the cluster stratified sampling design. Data were imputed with chained equations (40 imputations) in wide format, with covariates, PGIs, and BMI (z-scores) and diet MCA factor values included in imputation models. Estimates using multiply imputed data were pooled using Rubin’s (1987) rules.

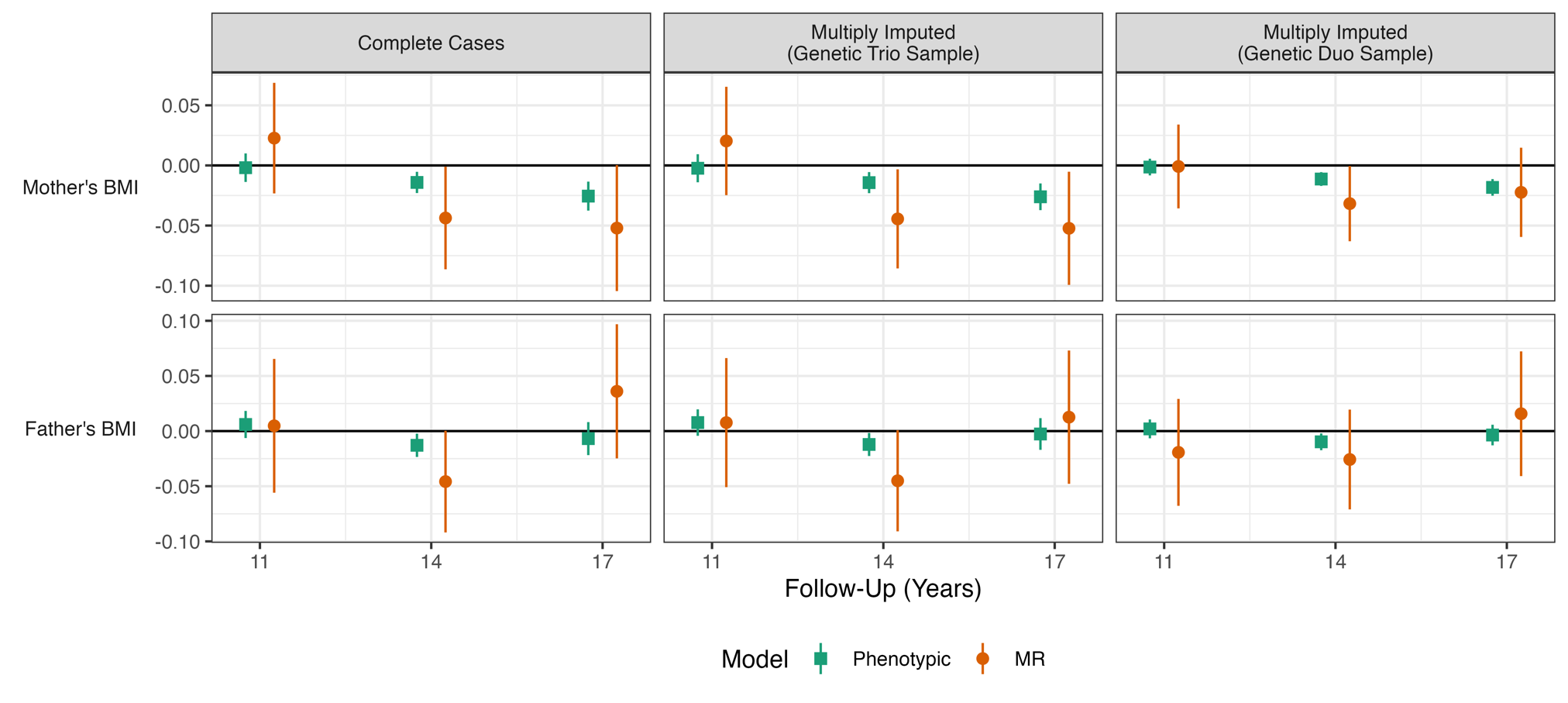

**Figure S8: Association between mother’s and father’s BMI and offspring diet by sample, PGIs used, and survey sweep.** Derived from Mendelian Rnadomization (MR; IV 2SLS) and ‘phenotypic’ multivariable regressions of offspring diet (MCA factor) on mother’s BMI, with adjustment for child’s PGI, sex, age at follow-up (two natural splines), maternal age at birth, family social class, mother’s education years, and 10 genetic principal components. In MR analysis, parental BMI instrumented with mother’s and father’s PGI. ‘Complete Cases’ refers to models using (outcome-sweep specific) complete cases data. ‘Multiply Imputed (Genetic Trio Sample)’ refers to models using multiply imputed data for the genotyped trio (mother-father-offspring) sample. ‘Multiply Imputed (Genetic Dup Sample)’ refers to models using multiply imputed data for the genotyped duo (mother-offspring or father-offspring) sample. All models also included adjustment for sex, age (two natural splines), maternal age at birth, family socioeconomic class, mother’s education years, and 10 genetic principal components. Regressions were weighted with recruitment weights accounting for the cluster stratified sampling design. Data were imputed with chained equations (40 imputations) in wide format, with covariates, PGIs, and BMI (z-scores) and diet MCA factor values included in imputation models. Estimates using multiply imputed data were pooled using Rubin’s (1987) rules.

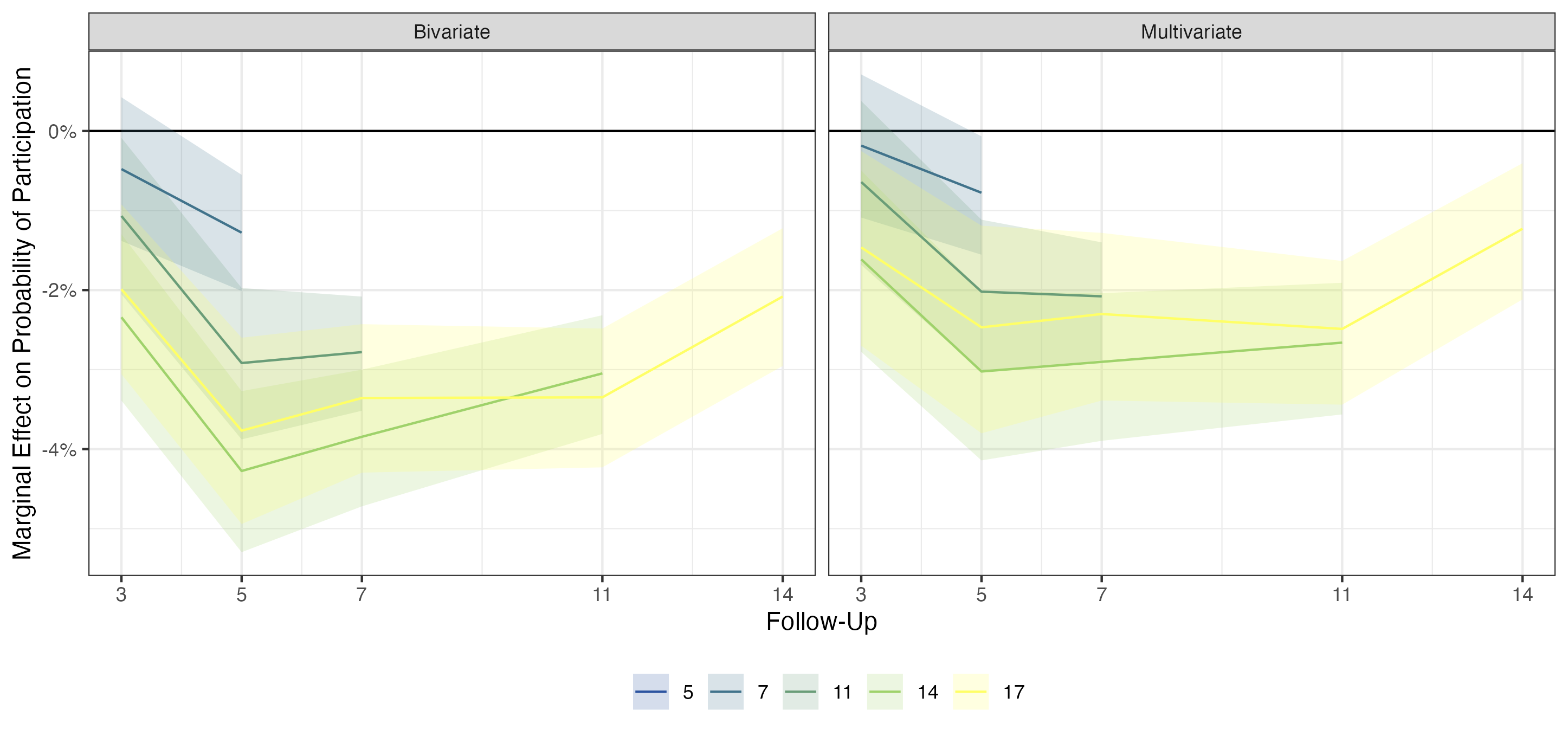

**Figure S9: Difference in probability of participation in given survey sweep according to BMI (z-score) at prior sweep.** Derived from logistic regression of participation on BMI (z-score) from a specific sweep. Colour of line indicates participation sweep, while location on x-axis indicates sweep relevant BMI (z-scores) collected. Left panel shows univariate regression, while right panel includes adjustment for sex, family socioeconomic class, maternal age at birth, mother’s BMI, father’s BMI, and mother’s years of education. Regressions weighted with recruitment weights accounting for the cluster stratified sampling design.

### Tables

**Table S1: Regression Results.** Association (+95% CI) between mother’s and father’s BMI and offspring diet and adiposity, by outcome variable, sweep, and estimator used. Genetic trio sample. ‘MR’ refers to Mendelian Randomization analysis using IV two stage least squares regression. In these models, parental BMI is instrumented with mother’s and father’s PGIs. ‘Phenotypic refers to multivariable regression analysis with parental (phenotypic) BMI entered into models directly (i.e. not instrumented with parental PGIs). ‘Z-Test’ refers to test examining differences in coefficients between MR and phenotypic models. Confidence intervals for this test were calculated using the Rao & Wu (1993) bootstrap method for clustered sampling designs (500 bootstraps, centile method; Kolenikov, 2010). Regressions were weighted with recruitment weights accounting for the cluster stratified sampling design and included adjustment for child’s PGI, sex, age (two natural splines), maternal age at birth, family socioeconomic class, mother’s education and years 10 genetic principal components (PCs).

|  | | Mother's BMI | | | Father's BMI | | |
| --- | --- | --- | --- | --- | --- | --- | --- |
| Outcome | Follow-Up | Phenotypic | MR | Z-Test | Phenotypic | MR | Z-Test |
| BMI (kg/m2) | 3y | 0.04 (0.03, 0.05) | 0.03 (-0.04, 0.11) | -0.01 (-0.09, 0.07) | 0.04 (0.03, 0.06) | -0.04 (-0.14, 0.07) | -0.07 (-0.18, 0.02) |
|  | 5y | 0.07 (0.05, 0.08) | 0.09 (0.01, 0.16) | 0.02 (-0.06, 0.10) | 0.07 (0.05, 0.09) | -0.03 (-0.12, 0.06) | -0.10 (-0.20, -0.02) |
|  | 7y | 0.09 (0.07, 0.11) | 0.10 (0.00, 0.20) | 0.01 (-0.09, 0.12) | 0.11 (0.09, 0.14) | -0.01 (-0.13, 0.10) | -0.12 (-0.24, -0.01) |
|  | 11y | 0.18 (0.15, 0.22) | 0.20 (0.06, 0.34) | 0.01 (-0.12, 0.16) | 0.21 (0.17, 0.25) | 0.05 (-0.13, 0.23) | -0.16 (-0.34, 0.03) |
|  | 14y | 0.22 (0.18, 0.26) | 0.16 (0.01, 0.31) | -0.06 (-0.23, 0.09) | 0.26 (0.22, 0.31) | 0.00 (-0.19, 0.20) | -0.25 (-0.47, -0.05) |
|  | 17y | 0.25 (0.20, 0.30) | 0.20 (-0.01, 0.42) | -0.05 (-0.27, 0.18) | 0.29 (0.24, 0.35) | -0.12 (-0.36, 0.13) | -0.41 (-0.69, -0.15) |
| Birthweight (g) | 0y | 14.53 (9.62, 19.45) | 23.04 (-0.75, 46.84) | 7.63 (-16.75, 34.44) | 1.85 (-3.55, 7.25) | -7.81 (-35.18, 19.57) | -8.35 (-40.16, 19.22) |
| BMI (Z-Score) | 3y | 0.03 (0.02, 0.04) | 0.03 (-0.02, 0.08) | 0.00 (-0.05, 0.05) | 0.03 (0.02, 0.04) | -0.02 (-0.09, 0.05) | -0.05 (-0.13, 0.01) |
|  | 5y | 0.04 (0.03, 0.05) | 0.06 (0.01, 0.11) | 0.02 (-0.03, 0.07) | 0.04 (0.03, 0.05) | -0.02 (-0.08, 0.03) | -0.06 (-0.12, -0.01) |
|  | 7y | 0.05 (0.04, 0.05) | 0.06 (0.01, 0.11) | 0.02 (-0.04, 0.06) | 0.06 (0.04, 0.07) | 0.00 (-0.05, 0.06) | -0.05 (-0.11, 0.01) |
|  | 11y | 0.06 (0.05, 0.07) | 0.07 (0.02, 0.13) | 0.01 (-0.05, 0.06) | 0.07 (0.06, 0.09) | 0.02 (-0.05, 0.08) | -0.06 (-0.12, 0.01) |
|  | 14y | 0.06 (0.05, 0.08) | 0.06 (0.01, 0.11) | -0.01 (-0.06, 0.04) | 0.08 (0.07, 0.09) | 0.00 (-0.06, 0.06) | -0.08 (-0.14, -0.02) |
|  | 17y | 0.07 (0.05, 0.08) | 0.07 (0.01, 0.13) | 0.00 (-0.06, 0.06) | 0.09 (0.07, 0.10) | -0.03 (-0.10, 0.04) | -0.12 (-0.19, -0.06) |
| Weight (Z-Score) | 3y | 0.02 (0.01, 0.03) | 0.03 (-0.02, 0.08) | 0.02 (-0.04, 0.07) | 0.03 (0.02, 0.05) | 0.00 (-0.06, 0.07) | -0.03 (-0.09, 0.03) |
|  | 5y | 0.03 (0.02, 0.04) | 0.05 (0.01, 0.10) | 0.02 (-0.03, 0.07) | 0.04 (0.03, 0.05) | -0.01 (-0.07, 0.05) | -0.05 (-0.12, 0.00) |
|  | 7y | 0.04 (0.03, 0.05) | 0.06 (0.01, 0.10) | 0.02 (-0.02, 0.06) | 0.05 (0.04, 0.06) | 0.00 (-0.05, 0.06) | -0.05 (-0.11, 0.01) |
|  | 11y | 0.05 (0.04, 0.06) | 0.06 (0.01, 0.11) | 0.01 (-0.04, 0.06) | 0.06 (0.05, 0.07) | 0.01 (-0.05, 0.07) | -0.05 (-0.12, 0.00) |
|  | 14y | 0.06 (0.05, 0.07) | 0.05 (0.01, 0.10) | 0.00 (-0.06, 0.04) | 0.07 (0.06, 0.09) | -0.01 (-0.07, 0.06) | -0.08 (-0.15, -0.02) |
|  | 17y | 0.06 (0.05, 0.07) | 0.07 (0.01, 0.13) | 0.01 (-0.07, 0.06) | 0.09 (0.07, 0.10) | -0.03 (-0.11, 0.04) | -0.12 (-0.20, -0.05) |
| Height (Z-Score) | 3y | 0.00 (-0.01, 0.01) | 0.02 (-0.03, 0.06) | 0.02 (-0.03, 0.07) | 0.02 (0.01, 0.03) | 0.02 (-0.04, 0.08) | 0.00 (-0.07, 0.04) |
|  | 5y | 0.00 (-0.01, 0.01) | 0.01 (-0.03, 0.05) | 0.01 (-0.03, 0.05) | 0.02 (0.01, 0.04) | 0.02 (-0.04, 0.07) | -0.01 (-0.06, 0.05) |
|  | 7y | 0.00 (-0.01, 0.01) | 0.02 (-0.02, 0.07) | 0.02 (-0.02, 0.07) | 0.02 (0.01, 0.03) | 0.00 (-0.05, 0.06) | -0.02 (-0.08, 0.03) |
|  | 11y | 0.01 (0.00, 0.02) | 0.01 (-0.03, 0.06) | 0.00 (-0.04, 0.05) | 0.02 (0.01, 0.04) | 0.00 (-0.06, 0.05) | -0.02 (-0.07, 0.03) |
|  | 14y | 0.01 (0.00, 0.02) | 0.02 (-0.03, 0.07) | 0.01 (-0.04, 0.06) | 0.01 (0.00, 0.02) | -0.02 (-0.07, 0.04) | -0.03 (-0.09, 0.03) |
|  | 17y | -0.01 (-0.02, 0.00) | 0.00 (-0.05, 0.05) | 0.01 (-0.04, 0.06) | 0.01 (0.00, 0.02) | 0.00 (-0.06, 0.06) | -0.01 (-0.07, 0.05) |
| Body Fat (%) | 7y | 0.17 (0.13, 0.22) | 0.16 (-0.07, 0.38) | -0.03 (-0.28, 0.20) | 0.20 (0.15, 0.26) | -0.03 (-0.27, 0.22) | -0.24 (-0.48, -0.01) |
|  | 11y | 0.37 (0.30, 0.44) | 0.30 (-0.03, 0.64) | -0.07 (-0.39, 0.28) | 0.41 (0.33, 0.50) | 0.27 (-0.09, 0.64) | -0.13 (-0.52, 0.22) |
|  | 14y | 0.39 (0.31, 0.46) | 0.20 (-0.10, 0.50) | -0.18 (-0.49, 0.08) | 0.46 (0.37, 0.54) | 0.10 (-0.26, 0.47) | -0.36 (-0.75, -0.03) |
|  | 17y | 0.40 (0.31, 0.49) | 0.17 (-0.20, 0.53) | -0.23 (-0.61, 0.11) | 0.46 (0.36, 0.57) | -0.17 (-0.59, 0.25) | -0.62 (-1.06, -0.21) |
| Fat-to-Non-Fat Ratio | 7y | 0.00 (0.00, 0.00) | 0.00 (0.00, 0.01) | 0.00 (0.00, 0.00) | 0.00 (0.00, 0.00) | 0.00 (-0.01, 0.00) | 0.00 (-0.01, 0.00) |
|  | 11y | 0.01 (0.01, 0.01) | 0.00 (0.00, 0.01) | 0.00 (-0.01, 0.00) | 0.01 (0.01, 0.01) | 0.00 (0.00, 0.01) | 0.00 (-0.01, 0.00) |
|  | 14y | 0.01 (0.01, 0.01) | 0.00 (0.00, 0.01) | 0.00 (-0.01, 0.00) | 0.01 (0.01, 0.01) | 0.00 (0.00, 0.01) | -0.01 (-0.01, 0.00) |
|  | 17y | 0.01 (0.01, 0.01) | 0.00 (0.00, 0.01) | 0.00 (-0.01, 0.00) | 0.01 (0.01, 0.01) | 0.00 (-0.01, 0.00) | -0.01 (-0.02, 0.00) |
| Fat Mass Index | 7y | 0.03 (0.02, 0.03) | 0.02 (-0.01, 0.06) | -0.01 (-0.04, 0.03) | 0.03 (0.02, 0.04) | 0.00 (-0.03, 0.04) | -0.03 (-0.07, 0.02) |
|  | 11y | 0.04 (0.03, 0.05) | 0.03 (0.00, 0.06) | -0.01 (-0.05, 0.02) | 0.04 (0.03, 0.05) | 0.02 (-0.02, 0.05) | -0.02 (-0.06, 0.02) |
|  | 14y | 0.08 (0.07, 0.10) | 0.05 (-0.01, 0.11) | -0.03 (-0.10, 0.02) | 0.09 (0.07, 0.11) | 0.02 (-0.06, 0.10) | -0.07 (-0.15, 0.01) |
|  | 17y | 0.10 (0.07, 0.12) | 0.10 (0.00, 0.19) | 0.00 (-0.10, 0.09) | 0.12 (0.09, 0.15) | -0.02 (-0.13, 0.08) | -0.14 (-0.25, -0.03) |
| Fat-Free Mass Index | 7y | 0.03 (0.02, 0.04) | 0.05 (0.01, 0.09) | 0.02 (-0.02, 0.06) | 0.04 (0.03, 0.05) | 0.02 (-0.02, 0.06) | -0.02 (-0.07, 0.03) |
|  | 11y | 0.05 (0.04, 0.05) | 0.06 (0.01, 0.10) | 0.01 (-0.03, 0.06) | 0.05 (0.04, 0.06) | -0.01 (-0.06, 0.04) | -0.06 (-0.12, -0.01) |
|  | 14y | 0.06 (0.05, 0.07) | 0.07 (0.02, 0.12) | 0.01 (-0.04, 0.06) | 0.08 (0.06, 0.10) | 0.00 (-0.07, 0.06) | -0.08 (-0.15, -0.02) |
|  | 17y | 0.09 (0.07, 0.11) | 0.14 (0.06, 0.23) | 0.05 (-0.04, 0.13) | 0.13 (0.10, 0.15) | -0.02 (-0.13, 0.08) | -0.14 (-0.26, -0.04) |
| Waist-to-Height Ratio | 5y | 0.00 (0.00, 0.00) | 0.00 (0.00, 0.00) | 0.00 (0.00, 0.00) | 0.00 (0.00, 0.00) | 0.00 (0.00, 0.00) | 0.00 (0.00, 0.00) |
|  | 7y | 0.00 (0.00, 0.00) | 0.00 (0.00, 0.00) | 0.00 (0.00, 0.00) | 0.00 (0.00, 0.00) | 0.00 (0.00, 0.00) | 0.00 (0.00, 0.00) |
| Diet Factor | 11y | 0.00 (-0.01, 0.01) | 0.02 (-0.02, 0.07) | 0.02 (-0.02, 0.07) | 0.01 (-0.01, 0.02) | 0.00 (-0.06, 0.07) | 0.00 (-0.06, 0.06) |
|  | 14y | -0.01 (-0.02, -0.01) | -0.04 (-0.09, 0.00) | -0.03 (-0.08, 0.01) | -0.01 (-0.02, 0.00) | -0.05 (-0.09, 0.00) | -0.03 (-0.08, 0.02) |
|  | 17y | -0.03 (-0.04, -0.01) | -0.05 (-0.10, 0.00) | -0.02 (-0.08, 0.03) | -0.01 (-0.02, 0.01) | 0.04 (-0.02, 0.10) | 0.04 (-0.03, 0.10) |
| Freq. Fast Food | 14y | 0.00 (-0.01, 0.00) | -0.03 (-0.05, 0.00) | -0.03 (-0.05, 0.00) | 0.00 (-0.01, 0.00) | -0.01 (-0.04, 0.01) | -0.01 (-0.04, 0.01) |
|  | 17y | -0.01 (-0.01, 0.00) | -0.02 (-0.05, 0.01) | -0.01 (-0.04, 0.01) | 0.00 (-0.01, 0.00) | 0.01 (-0.02, 0.04) | 0.01 (-0.02, 0.04) |
| Freq. Sugary Drinks | 11y | 0.00 (0.00, 0.01) | 0.01 (-0.01, 0.03) | 0.01 (-0.02, 0.03) | 0.00 (-0.01, 0.00) | 0.01 (-0.02, 0.03) | 0.01 (-0.02, 0.03) |
|  | 14y | 0.00 (0.00, 0.01) | 0.00 (-0.03, 0.02) | -0.01 (-0.03, 0.02) | 0.00 (-0.01, 0.00) | -0.01 (-0.03, 0.02) | -0.01 (-0.03, 0.02) |
|  | 17y | -0.01 (-0.01, 0.00) | -0.02 (-0.04, 0.01) | -0.01 (-0.04, 0.01) | 0.00 (-0.01, 0.01) | 0.01 (-0.02, 0.04) | 0.01 (-0.02, 0.04) |
| Freq. Sweetened Drinks | 11y | -0.01 (-0.02, -0.01) | -0.03 (-0.05, -0.01) | -0.02 (-0.04, 0.00) | 0.00 (-0.01, 0.00) | 0.00 (-0.02, 0.03) | 0.01 (-0.02, 0.03) |
|  | 14y | -0.01 (-0.02, -0.01) | -0.03 (-0.06, -0.01) | -0.02 (-0.05, 0.00) | -0.01 (-0.02, -0.01) | 0.00 (-0.03, 0.03) | 0.01 (-0.02, 0.04) |
|  | 17y | -0.01 (-0.02, -0.01) | -0.01 (-0.04, 0.02) | 0.00 (-0.03, 0.03) | -0.01 (-0.02, -0.01) | 0.02 (-0.01, 0.05) | 0.04 (0.00, 0.07) |
| Freq. Fruit | 14y | -0.01 (-0.01, 0.00) | -0.02 (-0.04, 0.00) | -0.01 (-0.03, 0.01) | 0.00 (-0.01, 0.00) | -0.02 (-0.05, 0.00) | -0.02 (-0.05, 0.01) |
|  | 17y | -0.01 (-0.02, -0.01) | -0.03 (-0.06, 0.00) | -0.02 (-0.05, 0.01) | 0.01 (0.00, 0.01) | 0.03 (0.00, 0.06) | 0.02 (-0.01, 0.06) |
| Freq. Vegetables | 14y | 0.00 (-0.01, 0.00) | -0.01 (-0.04, 0.01) | -0.01 (-0.03, 0.01) | 0.00 (-0.01, 0.00) | 0.00 (-0.03, 0.02) | 0.00 (-0.03, 0.02) |
|  | 17y | 0.00 (-0.01, 0.00) | -0.02 (-0.04, 0.01) | -0.01 (-0.04, 0.02) | 0.00 (-0.01, 0.01) | 0.00 (-0.03, 0.03) | 0.00 (-0.04, 0.03) |
| Daily Fruit or Vegetable | 3y | 0.00 (0.00, 0.00) | 0.00 (0.00, 0.01) | 0.00 (0.00, 0.02) | 0.00 (0.00, 0.00) | 0.00 (-0.01, 0.01) | 0.00 (-0.01, 0.01) |
| Portions Fruit per Day | 5y | 0.00 (0.00, 0.00) | -0.02 (-0.04, 0.01) | -0.02 (-0.04, 0.00) | 0.00 (0.00, 0.01) | 0.00 (-0.03, 0.03) | 0.00 (-0.03, 0.03) |
|  | 7y | 0.00 (-0.01, 0.00) | -0.01 (-0.03, 0.01) | -0.01 (-0.03, 0.01) | -0.01 (-0.01, 0.00) | -0.01 (-0.04, 0.02) | 0.00 (-0.03, 0.03) |
|  | 11y | 0.00 (-0.01, 0.00) | -0.03 (-0.05, 0.00) | -0.03 (-0.05, 0.00) | 0.00 (-0.01, 0.01) | 0.00 (-0.03, 0.03) | 0.00 (-0.03, 0.04) |

**Table S2: Regression Results.** Association (+95% CI) between child’s, father’s, or mother’s PGI and offspring BMI (z-score) or diet, by sweep. Genetic trio sample. Associations calculated using OLS regression. Child’s, father’s and mother’s PGIs entered into the models simultaneously. Child’s PGI reflects direct genetic effects and father’s and mother’s PGIs reflect indirect genetic effects. Regressions were weighted with recruitment weights accounting for the cluster stratified sampling design and included adjustment for sex, age (two natural splines), maternal age at birth, mother’s education years, and 10 genetic principal components (PCs).

|  | | PGI | | |
| --- | --- | --- | --- | --- |
| Outcome | Follow-Up | Child's | Mother's | Father's |
| BMI (kg/m2) | 3y | 0.11 (0.01, 0.20) | 0.03 (-0.06, 0.12) | -0.04 (-0.13, 0.06) |
|  | 5y | 0.17 (0.08, 0.27) | 0.10 (0.01, 0.19) | -0.03 (-0.11, 0.06) |
|  | 7y | 0.29 (0.15, 0.43) | 0.11 (-0.01, 0.23) | -0.01 (-0.12, 0.11) |
|  | 11y | 0.55 (0.36, 0.75) | 0.24 (0.06, 0.41) | 0.06 (-0.12, 0.23) |
|  | 14y | 0.71 (0.49, 0.94) | 0.19 (0.00, 0.37) | 0.02 (-0.17, 0.21) |
|  | 17y | 1.02 (0.72, 1.31) | 0.20 (-0.05, 0.45) | -0.10 (-0.34, 0.14) |
| Birthweight (g) | 0y | 9.26 (-22.89, 41.41) | 25.63 (-2.49, 53.76) | -5.55 (-30.82, 19.72) |
| BMI (Z-Score) | 3y | 0.07 (0.01, 0.14) | 0.02 (-0.03, 0.08) | -0.02 (-0.08, 0.04) |
|  | 5y | 0.11 (0.05, 0.17) | 0.07 (0.01, 0.12) | -0.02 (-0.07, 0.03) |
|  | 7y | 0.14 (0.08, 0.21) | 0.07 (0.01, 0.13) | 0.01 (-0.05, 0.06) |
|  | 11y | 0.21 (0.14, 0.28) | 0.09 (0.02, 0.15) | 0.02 (-0.04, 0.08) |
|  | 14y | 0.24 (0.17, 0.30) | 0.07 (0.01, 0.13) | 0.01 (-0.05, 0.07) |
|  | 17y | 0.30 (0.21, 0.38) | 0.07 (0.00, 0.14) | -0.02 (-0.08, 0.05) |
| Weight (Z-Score) | 3y | 0.04 (-0.03, 0.11) | 0.03 (-0.02, 0.09) | 0.00 (-0.06, 0.07) |
|  | 5y | 0.09 (0.03, 0.16) | 0.06 (0.00, 0.11) | -0.01 (-0.07, 0.05) |
|  | 7y | 0.11 (0.04, 0.17) | 0.06 (0.00, 0.12) | 0.01 (-0.05, 0.07) |
|  | 11y | 0.16 (0.10, 0.23) | 0.06 (0.00, 0.13) | 0.01 (-0.05, 0.07) |
|  | 14y | 0.19 (0.12, 0.26) | 0.06 (0.00, 0.12) | 0.00 (-0.06, 0.06) |
|  | 17y | 0.25 (0.17, 0.34) | 0.06 (-0.01, 0.14) | -0.02 (-0.09, 0.05) |
| Height (Z-Score) | 3y | -0.01 (-0.07, 0.06) | 0.02 (-0.03, 0.07) | 0.02 (-0.04, 0.07) |
|  | 5y | 0.03 (-0.03, 0.09) | 0.01 (-0.04, 0.06) | 0.01 (-0.04, 0.07) |
|  | 7y | 0.01 (-0.05, 0.08) | 0.02 (-0.03, 0.08) | 0.00 (-0.05, 0.06) |
|  | 11y | 0.03 (-0.03, 0.10) | 0.01 (-0.05, 0.07) | 0.00 (-0.06, 0.05) |
|  | 14y | 0.01 (-0.06, 0.08) | 0.02 (-0.04, 0.08) | -0.01 (-0.07, 0.05) |
|  | 17y | -0.03 (-0.10, 0.04) | 0.00 (-0.06, 0.05) | 0.00 (-0.06, 0.05) |
| Body Fat (%) | 7y | 0.60 (0.29, 0.92) | 0.19 (-0.09, 0.46) | 0.00 (-0.25, 0.25) |
|  | 11y | 1.00 (0.57, 1.43) | 0.40 (0.00, 0.80) | 0.29 (-0.07, 0.65) |
|  | 14y | 1.20 (0.78, 1.61) | 0.25 (-0.11, 0.62) | 0.12 (-0.24, 0.49) |
|  | 17y | 1.53 (1.03, 2.04) | 0.14 (-0.29, 0.58) | -0.11 (-0.51, 0.28) |
| Fat-to-Non-Fat Ratio | 7y | 0.01 (0.00, 0.02) | 0.00 (0.00, 0.01) | 0.00 (0.00, 0.00) |
|  | 11y | 0.02 (0.01, 0.02) | 0.01 (0.00, 0.01) | 0.01 (0.00, 0.01) |
|  | 14y | 0.02 (0.01, 0.03) | 0.00 (0.00, 0.01) | 0.00 (0.00, 0.01) |
|  | 17y | 0.03 (0.02, 0.03) | 0.00 (-0.01, 0.01) | 0.00 (-0.01, 0.01) |
| Fat Mass Index | 7y | 0.08 (0.03, 0.12) | 0.03 (-0.01, 0.07) | 0.01 (-0.03, 0.05) |
|  | 11y | 0.11 (0.07, 0.15) | 0.04 (0.00, 0.08) | 0.02 (-0.02, 0.05) |
|  | 14y | 0.26 (0.17, 0.35) | 0.06 (-0.02, 0.13) | 0.03 (-0.05, 0.10) |
|  | 17y | 0.36 (0.24, 0.49) | 0.11 (-0.01, 0.22) | 0.00 (-0.10, 0.10) |
| Fat-Free Mass Index | 7y | 0.08 (0.03, 0.13) | 0.06 (0.01, 0.10) | 0.02 (-0.02, 0.06) |
|  | 11y | 0.17 (0.12, 0.23) | 0.06 (0.01, 0.11) | -0.01 (-0.06, 0.04) |
|  | 14y | 0.22 (0.15, 0.29) | 0.08 (0.02, 0.14) | 0.00 (-0.06, 0.06) |
|  | 17y | 0.36 (0.24, 0.48) | 0.16 (0.05, 0.27) | 0.00 (-0.09, 0.10) |
| Waist-to-Height Ratio | 5y | 0.00 (0.00, 0.01) | 0.00 (0.00, 0.00) | 0.00 (0.00, 0.00) |
|  | 7y | 0.00 (0.00, 0.01) | 0.00 (0.00, 0.00) | 0.00 (0.00, 0.00) |
| Diet Factor | 11y | -0.02 (-0.08, 0.05) | 0.03 (-0.03, 0.08) | 0.01 (-0.05, 0.07) |
|  | 14y | 0.07 (0.01, 0.12) | -0.06 (-0.11, -0.01) | -0.05 (-0.09, 0.00) |
|  | 17y | -0.03 (-0.11, 0.04) | -0.06 (-0.12, 0.00) | 0.02 (-0.04, 0.08) |
| Freq. Fast Food | 14y | 0.04 (0.01, 0.07) | -0.04 (-0.06, -0.01) | -0.02 (-0.04, 0.00) |
|  | 17y | 0.02 (-0.02, 0.05) | -0.03 (-0.06, 0.00) | 0.00 (-0.03, 0.03) |
| Freq. Sugary Drinks | 11y | -0.01 (-0.04, 0.02) | 0.01 (-0.02, 0.04) | 0.01 (-0.02, 0.03) |
|  | 14y | 0.01 (-0.02, 0.04) | -0.01 (-0.03, 0.02) | -0.01 (-0.03, 0.02) |
|  | 17y | 0.00 (-0.04, 0.04) | -0.02 (-0.05, 0.01) | 0.00 (-0.03, 0.03) |
| Freq. Sweetened Drinks | 11y | 0.01 (-0.02, 0.05) | -0.03 (-0.06, -0.01) | 0.00 (-0.02, 0.03) |
|  | 14y | -0.01 (-0.04, 0.02) | -0.04 (-0.06, -0.01) | 0.00 (-0.03, 0.02) |
|  | 17y | -0.03 (-0.07, 0.01) | -0.01 (-0.05, 0.02) | 0.02 (-0.01, 0.05) |
| Freq. Fruit | 14y | 0.03 (0.01, 0.06) | -0.02 (-0.05, 0.00) | -0.02 (-0.05, 0.00) |
|  | 17y | -0.01 (-0.04, 0.03) | -0.03 (-0.06, 0.00) | 0.02 (-0.01, 0.05) |
| Freq. Vegetables | 14y | 0.01 (-0.02, 0.04) | -0.02 (-0.04, 0.01) | 0.00 (-0.03, 0.02) |
|  | 17y | 0.00 (-0.04, 0.03) | -0.02 (-0.05, 0.02) | -0.01 (-0.04, 0.02) |
| Daily Fruit or Vegetable | 3y | -0.01 (-0.02, 0.00) | 0.01 (0.00, 0.02) | 0.00 (-0.01, 0.01) |
| Portions Fruit per Day | 5y | 0.00 (-0.03, 0.03) | -0.02 (-0.04, 0.01) | 0.00 (-0.03, 0.02) |
|  | 7y | 0.01 (-0.02, 0.03) | -0.01 (-0.03, 0.02) | -0.01 (-0.03, 0.02) |
|  | 11y | -0.01 (-0.04, 0.03) | -0.03 (-0.06, 0.00) | 0.00 (-0.03, 0.03) |

**Table S3: Descriptive statistics by sample.** ‘All’ refers to the full eligible sample (n = 15,180); ‘Genetic Trios’ to the sample with genotyped mother-father-offspring triads (n = 2,562); and ‘Mother-Offspring Pairs’ to the sample with genotyped mothers and offspring (n = 5,184). Descriptive statistics are weighted with recruitment weights accounting for the cluster stratified sampling design. Wald tests for each variable were carried out to assess whether there were mean level differences between ‘Genetic Trios’ and non-Genetic Trios and Mother-Offspring Pairs and non-Mother-Offspring Pairs. *** p < 0.01; ** p < 0.01; * p < 0.01.

|  | | Eligible Sample | | Genetic Trios | | Mother-Offspring Pairs | |
| --- | --- | --- | --- | --- | --- | --- | --- |
|  | Variable | Mean (SD) / n (%) | Missing % | Mean (SD) / n (%) | Missing % | Mean (SD) / n (%) | Missing % |
|  | Birthweight (g) | 3424.7 (519.1) | 1.6% | 3481.4 (504.2)*** | 0.9% | 3445.5 (514)** | 1.1% |
|  | BMI (Z-Score) @ 3y | 0.5 (0.9) | 25.9% | 0.5 (0.9) | 12.8% | 0.5 (0.9) | 14.5% |
|  | BMI (Z-Score) @ 5y | 0.5 (0.9) | 21.8% | 0.4 (0.9) | 5% | 0.5 (0.9) | 6.4% |
|  | BMI (Z-Score) @ 7y | 0.4 (1) | 28.2% | 0.3 (1) | 4.8% | 0.4 (1) | 7.7% |
|  | BMI (Z-Score) @ 11y | 0.5 (1.2) | 32.8% | 0.4 (1.1)** | 3.5% | 0.5 (1.2) | 5.8% |
|  | BMI (Z-Score) @ 14y | 0.5 (1.2) | 43.3% | 0.5 (1.1) | 3.1% | 0.6 (1.2)** | 3.5% |
|  | BMI (Z-Score) @ 17y | 0.5 (1.2) | 52.3% | 0.5 (1.2) | 16.4% | 0.6 (1.2)* | 22.8% |
|  | Body Fat (%) @ 7y | 20.6 (4.7) | 29.5% | 20.4 (4.7) | 6.2% | 20.6 (4.6) | 9.3% |
|  | Body Fat (%) @ 11y | 21.7 (7.4) | 33.4% | 21.3 (7.2)** | 4% | 21.8 (7.4) | 6.6% |
|  | Body Fat (%) @ 14y | 21.2 (8.9) | 43.7% | 21 (8.7) | 3% | 21.5 (8.9)** | 3.7% |
|  | Body Fat (%) @ 17y | 21.6 (10) | 52.8% | 21.5 (9.5) | 16.9% | 21.9 (10)* | 23.5% |
|  | Fat-to-Non-Fat Ratio @ 7y | 0.3 (0.1) | 29.4% | 0.3 (0.1) | 6.2% | 0.3 (0.1) | 9.3% |
|  | Fat-to-Non-Fat Ratio @ 11y | 0.3 (0.1) | 33.7% | 0.3 (0.1)** | 4.5% | 0.3 (0.1) | 7.1% |
|  | Fat-to-Non-Fat Ratio @ 14y | 0.3 (0.1) | 44.1% | 0.3 (0.1) | 3.7% | 0.3 (0.2)** | 4.5% |
|  | Fat-to-Non-Fat Ratio @ 17y | 0.3 (0.2) | 53.3% | 0.3 (0.2) | 17.5% | 0.3 (0.2)** | 24.4% |
|  | Waist-to-Height Ratio @ 5y | 0.5 (0) | 22.1% | 0.5 (0) | 5% | 0.5 (0) | 6.5% |
|  | Waist-to-Height Ratio @ 7y | 0.5 (0) | 29% | 0.5 (0)* | 5.3% | 0.5 (0) | 8.4% |
|  | Diet Factor @ 11y | 0 (1) | 30.8% | 0 (1)* | 1.6% | 0 (1) | 3.8% |
|  | Diet Factor @ 14y | -0.1 (1) | 41.4% | 0 (1)*** | 1.2% | -0.2 (1) | 1.2% |
|  | Diet Factor @ 17y | -0.1 (1) | 64.7% | 0 (1)*** | 35% | -0.1 (1) | 41.7% |
| Sex | Male | 7,949.7 (51.4%) | 0% | 1,322.6 (50.3%) | 0% | 2,687.4 (50.1%)* | 0% |
|  | Female | 7,506.3 (48.6%) |  | 1,306.4 (49.7%) |  | 2,680.6 (49.9%) |  |
|  | Maternal Age at Birth | 29.4 (5.9) | 0.5% | 31.4 (4.8)*** | 0.3% | 30.1 (5.5)*** | 0.4% |
|  | Mother's BMI | 24.8 (4.8) | 3.2% | 25.1 (4.8)** | 1.2% | 25 (4.9)*** | 1.4% |
|  | Father's BMI | 26.1 (3.9) | 21.9% | 26.3 (3.7)* | 1.8% | 26.3 (3.8)** | 14.9% |
| Family Social Class (NS-SEC) | Managerial and Professional | 6,731.0 (44.7%) | 2.6% | 1,672.6 (63.6%)*** | 0% | 2,824.9 (52.6%)*** | 0% |
|  | Intermediate | 1,427.9 (9.5%) |  | 259.1 (9.9%) |  | 536.9 (10%) |  |
|  | Small Employer and Self-Employed | 1,118.2 (7.4%) |  | 184.3 (7%) |  | 377.6 (7%) |  |
|  | Lower Supervisory & Technical | 1,261.0 (8.4%) |  | 197.4 (7.5%) |  | 425.3 (7.9%) |  |
|  | Semi-Routine and Routine | 1,886.8 (12.5%) |  | 213.2 (8.1%) |  | 597.4 (11.1%) |  |
|  | Not Working | 2,636.9 (17.5%) |  | 102.5 (3.9%) |  | 604.0 (11.3%) |  |
|  | Mother's Years of Education | 6.2 (1.7) | 0.7% | 6.8 (1.8)*** | 0.2% | 6.5 (1.8)*** | 0.4% |
|  | Father's Years of Education | 6.1 (1.8) | 20.7% | 6.4 (1.9)*** | 1.6% | 6.2 (1.9)*** | 14.1% |
| Country of Birth | England | 12,440.9 (80.5%) | 0% | 2,133.1 (81.1%) | 0% | 4,379.7 (81.6%)* | 0% |
|  | Wales | 855.4 (5.5%) |  | 146.0 (5.6%) |  | 301.7 (5.6%) |  |
|  | Scotland | 1,573.5 (10.2%) |  | 251.0 (9.5%) |  | 494.7 (9.2%) |  |
|  | Northern Ireland | 586.3 (3.8%) |  | 98.9 (3.8%) |  | 191.9 (3.6%) |  |

**Table S4: Description of individual diet variables.**

| Variable | Survey Question | Labels | Variables | 3y | 5y | 7y | 11y | 14y | 17y |
| --- | --- | --- | --- | --- | --- | --- | --- | --- | --- |
| Daily Fruit or Vegetable | I would now like to ask you about things that relate to people's standard of living. Do you have any of the following items?....Fresh fruit or vegetables once a day for [child] | Yes; No | bpstfv00 | x |  |  |  |  |  |
| Portions Fruit per Day | On a typical day, how many portions of fresh, frozen, tinned or dried fruit does [child] eat? | None; One; Two; Three or more | cpfrtp00, dpfrtp00, epfrtp00 |  | x | x | x |  |  |
| Freq. Sugary Drinks | Parent Report: How often, if at all, does [child] drink sweetened drinks e.g. cola, squash or Sunny Delight?  Child Report: How often, if at all, do you drink sugary drinks like regular cola or squash? | More than once a day; Once a day; 3-6 days a week; 1-2 days a week; Less often but at least once a month; Less than once a month; Never; Don't know; Refused | epswtd00, fcswtd00, gcswtd00 |  |  |  | x | x | x |
| Freq. Sweetened Drinks | Parent Report: How often, if at all, does [child] drink artificially sweetened drinks e.g. diet cola, sugar-free squash?  Child Report: How often, if at all, do you drink diet drinks or sugar free drinks like diet cola or sugar-free squash? | More than once a day; Once a day; 3-6 days a week; 1-2 days a week; Less often but at least once a month; Less than once a month; Never; Don't know; Refused | epaswd00, fcaswd00, gcaswd00 |  |  |  | x | x | x |
| Freq. Fast Food | How often, if at all, do you eat fast food such as McDonalds, Burger King, KFC or other fast food like that? | More than once a day; Once a day; 3-6 days a week; 1-2 days a week; Less often but at least once a month; Less than once a month; Never | fctkwy00, gctkwy00 |  |  |  |  | x | x |
| Freq. Fruit | How often do you eat at least 2 portions of fruit per day?  A portion of fruit could be a whole piece of fruit, like an apple or banana or 80g of fruit (like in a fruit salad) but does not include fruit juices. | Never; Some days, but not all days; Every day | fcfrut00, gcfrut00 |  |  |  |  | x | x |
| Freq. Vegetables | How often do you eat at least 2 portions of vegetables including salad, fresh, frozen or tinned vegetables per day?  A portion is 3 heaped tablespoons of cooked vegetables or beans /pulses or a handful of cherry tomatoes or a small bowl of salad. It does not include potatoes. | Never; Some days, but not all days; Every day | fcvegi00, gcvegi00 |  |  |  |  | x | x |
